# Identifying selective predictors of ADHD, Oppositional Defiant and Conduct Disorder onset in early adolescence with optimized deep learning

**DOI:** 10.1101/2023.08.19.23294322

**Authors:** Nina de Lacy, Michael J. Ramshaw

## Abstract

**Introduction:** The externalizing disorders of ADHD, Oppositional Defiant Disorder (ODD) and Conduct Disorder (CD) exhibit a strong uptick in incidence in late childhood to become some of the most common mental health conditions in adolescence and strong predictors of adult psychopathology. While treatable, substantial diagnostic overlap exists among the externalizing disorders, complicating intervention planning. Thus, early adolescence is a period of considerable interest in understanding which factors predict the onset of externalizing disorders and disambiguating those that may differentially predict the development of ADD versus (vs) ODD and CD.

**Materials and Methods:** Here, we analyzed 5,777 multimodal candidate predictors collected from children age 9-10 yrs and their parents in the ABCD cohort spanning demographics; developmental and medical history; physiologic function; academic performance; social, physical and cultural environment; activities of everyday life, substance use and cortical and subcortical brain structure, volumetrics, connectivity and function to predict the future onset of ADHD, ODD and CD at 2-year follow-up. We used deep learning optimized with an innovative AI algorithm that jointly optimizes model training and performs automated feature selection to construct prospective, individual-level predictions of illness onset in this high-dimension data. Additional experiments furnished predictive models of all prevailing cases at 11-12 yrs and examined relative predictive performance when candidate predictors were restricted to only neural metrics derived from MRI.

**Results:** Multimodal models achieved strong, consistent performance with ∼86-97% accuracy, 0.919-0.996 AUROC and ∼82-97% precision and recall in testing in held-out, unseen data. In neural-only models, predictive performance dropped substantially but nonetheless accuracy and AUROC of ∼80% were achieved. Parent aggressive and externalizing traits uniquely differentiated the onset of ODD while structural MRI metrics in the limbic system specifically predicted the onset of CD. Psychosocial measures of sleep disorders, parent mental health and behavioral traits and school performance proved valuable across all disorders but cognitive and non-neural physiologic metrics were never selected. In neural-only models, structural and functional MRI metrics in subcortical regions and cortical-subcortical connectivity were emphasized over task fMRI or diffusion measures. Overall, we identified a strong correlation between accuracy and final predictor importance.

**Conclusions:** Deep learning optimized with AI can generate highly accurate individual-level predictions of the onset of early adolescent externalizing disorders using multimodal features. Analysis of 5,777 multimodal candidate predictors highlighted psychosocial predictors related to sleep disorders, school performance and parent mental health and behavioral traits over other feature types. While externalizing disorders are frequently co-morbid in adolescents, certain predictors appeared specific to the onset of ODD or CD vs ADHD with structural MRI metrics in the limbic system offering particular promise in identifying children at risk for the onset of CD, a highly disabling disorder. The strong observed correlation between predictive accuracy and final predictor importance suggests that principled, data-driven searches for impactful predictors may facilitate the construction of robust, individual-level models in high-dimension data. To our knowledge, this is the first machine learning study to predict the onset of all three major adolescent externalizing disorders with the same design and participant cohort to enable direct comparisons, analyze >200 multimodal features and include as many types of neuroimaging metrics. Future work to test our observations in external validation data will help further test the generalizability of these findings.

## INTRODUCTION

Attention Deficit Hyperactivity Disorder (ADHD), Oppositional Defiant Disorder (ODD) and Conduct Disorder (CD) are common mental health conditions in adolescence, often collectively referred to as the externalizing disorders. Among the most common of youth mental health conditions, externalizing behaviors are the most frequent reason for referral to mental health services and a strong predictor of adult psychopathology. (1) In school age youth (K-12), 10-24% meet criteria for externalizing disorders, with ADHD and ODD being the most common. (2) ADHD affects 7-10% of youth < 18 years of age (yrs) with prevalence showing a strong uptick in early adolescence, peak in mid-late adolescence and decline into adulthood. Some ∼2% of children ≤5 yrs are affected versus (vs) ∼10% at 6-11 yrs and ∼13% at 12-17 yrs, with ∼4% of adults having clinical ADHD (2, 3, 4). In contrast, ODD and CD (collectively the disruptive externalizing disorders) affect ∼5% of youth ≤17 yrs growing to ∼10-12% of adults, where in the latter they are associated with increased risk for later co-morbid mental health and substance use disorders (5, 6, 7, 8). Among youth with ADHD, ∼30-50% may also exhibit disruptive externalizing behaviors consistent with ODD and CD, with this association growing with increasing age and linked to later poor academic and life outcomes such as school dropout, substance abuse and involvement with the justice system. (9, 10, 11, 12, 13). Thus, early adolescence is a period of considerable interest in understanding which risk factors predict the onset of externalizing disorders and disambiguating those that may differentially predict the development of ADD vs ODD and CD.

Adolescent externalizing disorders have attracted a range of research approaches. Historically, these have predominantly been cross-sectional studies quantifying group-level associations, frequently assessing neuroimaging metrics. More recently, machine learning (ML) classification techniques have been applied, increasingly to large-scale datasets. Such approaches offer the advantages of providing individual-level case predictions from high dimension and/or multimodal data, thereby bridging from extant work focused on identifying statistical associations at a group level to a pathway toward personalized medicine. (14, 15) Appropriately constructed, ML algorithms can simultaneously analyze hundreds or thousands of candidate predictors and enlarge the solution space. Such work has been further fueled by the increasing availability of large-scale, open science datasets incorporating multimodal variables. In peri-adolescence, the flagship initiative of this type is the ongoing population-level, longitudinal ABCD study (*n*=11,800) used in the present study that enrolled children at age 9-10 yrs and collects data from many knowledge domains including multiple neuroimaging types (16, 17, 18). While a number of ML predictive studies have been performed in adolescent externalizing disorders, these have to date largely (though not exclusively) been cross-sectional and focused on predicting prevailing cases at a particular age in a single disorder. Few ML studies have predicted the future onset of disease in longitudinal data or applied a consistent analytic architecture across the three major adolescent externalizing disorders in the same population cohort to enable direct comparisons.

In the present study, we extend prior work with an ML design that analyzes a large number of multidomain candidate predictors to predict new onset cases of ADHD, ODD and CD in early adolescence in the same design and youth cohort. We aimed to identify the best-performing predictors and compare these across these three related disorders to understand whether there were shared or unique predictors underpinning ADHD, ODD and CD. Given the large prior literature related to brain structure and function motifs in the externalizing disorders, we also wanted to compare the relative predictive ability of models composed purely of neuroimaging metrics derived from MRI with multimodal models. By leveraging an AI algorithm that jointly optimizes ML model training and performs automated feature selection, we were able to analyze 5,777 candidate predictors spanning demographics; developmental and medical history; white and gray matter brain structure, neural function (cortical and subcortical connectivity, 3 tasks); brain volumetrics; physiologic function (e.g. sleep, hormone levels, pubertal stage, physical function); cognitive and academic performance; social and cultural environment (e.g. parents, friends, bullying); activities of everyday life (e.g. screen use, hobbies); living environment (e.g. crime, pollution, educational and food availability) and substance use. We used features assessed at 9-10 yrs (107-132 months) to predict future, new onset cases of ADHD, ODD and CD at 11-12 years with deep learning with artificial neural networks, which incorporates non-linear relationships among predictors and is resistant to multicollinearity. Since extant work is more focused on predicting prevailing rather than new onset cases, we performed additional experiments to predict all prevailing cases at 11-12 yrs to provide comparisons with the existing literature. Our AI approach allowed us to render fully interpretable predictions, quantify relative predictor importance at both the group- and individual-level and examine the relationship between model accuracy and predictor importance across all models. All results presented are from testing for generalization in holdout, unseen data.

## MATERIALS AND METHODS

### Terminology and definitions

Terms used in quantitative analysis may be shared among different fields with variant meanings. Here, we use ML conventions throughout. (26, 27, 28) ‘Prediction’ means predicting the quantitative value of a target variable by analyzing patterns in input data. The set of observations used to train and validate models is referred to as the ‘training set’ and the unseen holdout set of observations is termed the ‘test set’. We refer to the set of all input data used in training as containing ‘features’ or ‘candidate predictors’ and those identified in final, optimized models after testing in held-out data (presented in **Results)** as ‘final predictors’. We use ‘generalizability’ to refer to the ability of a trained model to adapt to new, previously unseen data drawn from the same distribution i.e. model fit in the test set. ‘Precision’ refers to the fraction of positive predictions that were correct; ‘Recall’ to the proportion of true positives that were correctly predicted; and ‘Accuracy’ to the number of correct predictions as a fraction of total predictions. Receiver Operating Characteristic curves (ROC Curves) are provided that quantify classification performance at different classification thresholds plotting true positive versus false positive rates, where the Area Under the Curve (AUROC) is defined as the two-dimensional area under the ROC curve from (0,0) to (1,1). This paragraph defining terminology usage is adapted from our prior work.

### Data and data collection in the ABCD study

We use data from the ABCD study, an epidemiologically-informed prospective cohort study that recruited 11,880 children (52% male; 48% female) aged 9-10 years in 21 sites across the United States, intending to follow this youth for the next decade. Participants in the cohort include 800 twin pairs (*n*=800) and/or non-twin siblings. This data is made available to qualified researchers at no cost from the National Institute of Mental Health Data Archive and is released periodically. The present study uses data from Release 4.0, the 42-month follow-up date. Fuller descriptions of the overall design of the ABCD study as well as recruitment procedures and the participant sample may be found in Jernigan et al; Garavan et al; and Volkow et al. (29, 30, 31) This study has been reviewed and deemed not human subjects research by the University of Utah Institutional Review Board.

ABCD collects a wide range of information from youth participants and their parents comprising phenotypic, demographic, psychometric, physiologic and developmental data as well as multiple modalities of MRI neuroimaging. Barch et al and Lisdahl et al respectively detail the phenotypic and substance abuse assessment protocols. (32, 33) Here, we utilize data from assessments of physical and mental health, substance use, neurocognition, school performance and quality, culture and environment performed for youth and their parents as well as biospecimens (e.g. pubertal hormone levels) and environmental toxin exposure. A summary description of assessments performed and environmental and school-related variables derived from geocoding at age 9-10 yrs that we analyzed may be inspected in **Supplementary Table 1**.

Brain imaging incorporates optimized 3D T1; 3D T2; Diffusion Tensor Imaging; Resting state functional MRI (rsfMRI); and 3 task MRI (tfMRI) protocols harmonized across acquisition sites. The tfMRI protocol comprises the Monetary Incentive Delay (MID) and Stop Signal (SST) tasks and an emotional version of the n-back task which collectively measure reward processing, motivation, impulsivity, impulse control, working memory and emotion regulation. In the present study we utilized ABCD-provided fully-processed metrics from each of these imaging types that are computed after quality control. Detailed descriptions of the requisite acquisition, pre-processing, quality control and analytic protocols used to generate metrics may be inspected in Casey et al and Hagler et al. (34, 35) We utilize all available processed metrics that have passed quality control from diffusion fullshell; cortical and subcortical Gordon correlations (connectivity); structural; volumetric; and all three tfMRI tasks as well as corresponding head motion statistics for each modality. For certain modalities such as rsfMRI, multiple scans were attempted or completed. In such cases we use metrics computed from the first scan.

### Study inclusion criteria and sample partitioning for machine learning

Inclusion criteria for the present study were a) participants enrolled in the study at baseline (9-10 yrs) who were still enrolled in the ABCD study at 2-year follow-up at 11-12 yrs (*n*=8,085) who had b) complete data for all neural imaging types for at least one scan in each modality listed above that passed ABCD quality control (*n*=6,178) and were c) youth participants unrelated to any other youth participant in the study (*n*=5,355). If a youth had sibling(s) present in the cohort, we selected the oldest sibling for inclusion. Demographic characteristics of this sample at age 9-10 yrs, the age which corresponds to input data used to make predictions, is presented below in **Table 1**.

**Table 1:** Demographic characteristics of participant sample at age 9-10 years. Sex refers to sex assigned at birth on the original birth certificate. Gender refers to the youth’s gender identification. Race and ethnicity refer to the parents’ view of youth’s race or ethnicity. More than one race or ethnicity identification may be selected and therefore percentages may sum to >100%.

| Characteristic | Number | Percent |
| --- | --- | --- |
| **Sex** |  |  |
| Male | 2,771 | 51.7% |
| Female | 2,584 | 48.3 |
| **Gender Identity** |  |  |
| Male | 2,768 | 51.7% |
| Female | 2,577 | 48.1 |
| Gender non-conforming | 7 | 0.1 |
| Don't know/didn't answer | 4 | 0.1 |
| **Race** |  |  |
| Black/African American | 873 | 16.3% |
| Asian | 353 | 6.6 |
| White | 4,236 | 79.1 |
| Native American/Alaska Native | 187 | 3.5 |
| Other | 334 | 6.2 |
| **Ethnicity** |  |  |
| Hispanic/Latino/Latinx | 1,070 | 20.0% |
| Non-Hispanic | 4,224 | 78.9 |
| Not indicated | 62 | 1.2 |

Physiologic and cognitive characteristics of the same participant sample at 9-10 yrs may be viewed in **Table 2**.

**Table 2:** Physiologic and cognitive characteristics of participant sample at age 9-10 years. Characteristics of the study sample at 9-10 yrs. Pubertal development is measured with the Pubertal Development Scale (adapted from the Petersen scale) in a sex-specific manner. Height is measured twice with the average of these values presented. We note a range of 11.0-255.0 pounds for weight which is the range present in the original ABCD data. Handedness is assessed with the Edinburgh Handedness Inventory. Cognitive metrics are assessed with the NIH Toolbox and are all age-corrected scores. Vocabulary is measured with the Picture Vocabulary test; Attention and inhibition with the Flanker Inhibitory Control & Attention Task; Executive Function with the Dimensional Change Card Sort Test; and Processing Speed with the Pattern Comparison Processing Speed Test.

| Characteristic | Range | Mean | Median |
| --- | --- | --- | --- |
| **Age in months** | 107.0-132.0 | 119.9 | 120.0 |
| **Pubertal Development Stage** | 1-5 | 2 | 2 |
| **Height (inches)** | 36.6-81.0 | 55.4 | 55.4 |
| **Weight (pounds)** | 11.0-255.0 | 82.3 | 76.8 |
| **Waist Circumference (cm)** | 17.0-61.0 | 26.4 | 25.5 |
| **Handedness** |  |  |  |
| Writing | -100.0-100.0 | 76.5 | 100.0 |
| Throwing | -100.0-100.0 | 67.1 | 100.0 |
| Spoon | -100.0-100.0 | 62.4 | 100.0 |
| **Vocabulary** | 51.0-208.0 | 109.0 | 109.0 |
| **Attention and Inhibition** | 65.0-171.0 | 96.5 | 97.0 |
| **Working Memory** | 46.0-194.0 | 102.0 | 103.0 |
| **Executive Function** | 68.0-181.0 | 98.0 | 94.0 |
| **Processing Speed** | 20.0-185.0 | 95.2 | 95.0 |

The final participant sample (n=5,356 participants) after inclusion criteria were applied was randomly partitioned into a training set comprising 70% of the sample (*n*=3,749) and a holdout, unseen test set comprising 30% of the sample (*n*=1,607, **Figure 1**). This partitioning was effected prior to pre-processing either input features (candidate predictors) or predictive target to minimize bias.

**Figure 1:**
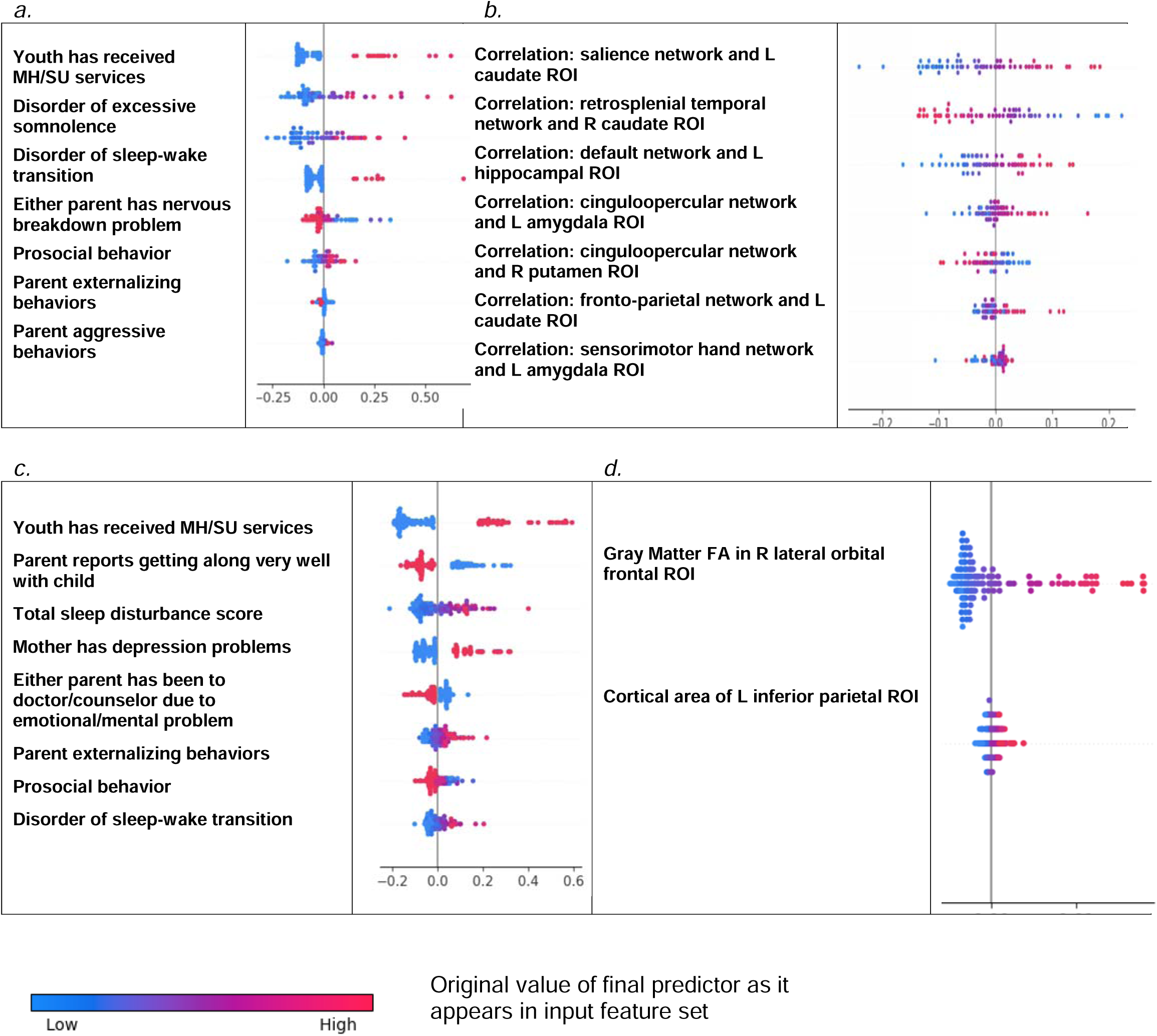
Formation of the study participant sample. Steps used to form the study sample are shown. After inclusion criteria are applied, the sample was randomly partitioned into training and test sets followed by separate pre-processing of targets and features. Subsequently, samples for each experiment were formed as described in **Preparation of predictive targets** and **Construction of participant samples for cases with externalizing disorders and controls**

### Preparation of predictive targets

Predictive targets of ADHD, ODD and CD cases were derived from the Child Behavior Checklist for youth ages 4-18 years (CBCL) known as the ‘ABCD Parent Child Behavior Checklist Scores Aseba (CBCL)’ in ABCD study nomenclature. The CBCL is a standardized instrument in widespread clinical and research use. It forms part of the Achenbach System of Empirically Based Assessment (ASEBA) “designed to facilitate assessment, intervention planning and outcome evaluation among school, mental health, medical and social service practitioners who deal with maladaptive behavior in children, adolescents and young adults.” (36) To score the CBCL, parents rate their child on a 0-1-2 scale on 118 specific problem items such as “Acts too young for age” over the prior 6 months. Answers are aggregated into raw, T and percentile scores for 8 syndrome subscales (Anxiety, Somatic Problems, Depression, Social Problems, Thought Problems, Attention Problems, Rule Breaking and Aggressive Behavior) derived from principal components analysis of data from 4455 children referred for mental health services. The CBCL is normed in a U.S. nationally representative sample of 2368 youth ages 4-18 yrs that takes into account differences in problem scores for “males versus females”. It exhibits excellent test-retest reliability of 0.82-0.96 for the syndrome scales with an average *r* of 0.89 across all scales. Content and criterion validity is strong with referred versus non-referred children scoring higher on 113/188 problem items and significantly higher on all problem scales, respectively.

To form binary classification targets, we thresholded CBCL subscale T scores for ADHD (“attention problems”), ODD (“aggressive behavior”) and CD (“rule breaking”) using cutpoints established by ASEBA for clinical practice. Specifically, a T score of 65-69 (95^th^ to 98^th^ percentile) is considered in the ‘borderline clinical’ range, and scores of ≥70 are considered in the ‘clinical range.’ Accordingly, we discretized T scores for each of the 3 subscales under consideration by deeming every individual with a T score ≥ 65 as a ‘case’ or [1] and every individual with a score <65 as a ‘not case’ or [0]. This process was performed separately in the training and test sets for participant CBCL scores at 9-10 yrs and at 11-12 yrs.

### Construction of participant samples for cases with externalizing disorders and controls

We formed two participant samples in each of ADHD, ODD and CD in the training and test sets (**Figure 1**). The first sample type comprised all cases of ADHD, ODD and CD present in the larger sample at 11-12 yrs. The second sample type comprised only new onset cases at 11-12 yrs. A new onset case was defined as a youth who met criteria for ADHD, ODD or CD as defined by the ASEBA CBCL cut-points at 11-12 yrs who did not meet criteria for the requisite disorder at 9-10 yrs. Thus, six participant samples in total were constructed. In all samples, we formed a balanced sample of cases and controls. The latter were youth with the lowest possible scores on the relevant syndrome scale selected from the eligible study population (see: **Baseline inclusion criteria and sample partitioning for machine learning**) and matched with cases for age in months and sex/gender.

### Preparation of candidate predictors (input features)

We assembled a feature set for input into predictive algorithms that comprised the majority of phenotypic, demographic, psychometric, physiologic and developmental variables available from the ABCD study (including data collection site) and all available neural metrics including head motion statistics with the exception of temporal variance measures (**Supplementary Table 1**). We used only metrics collected at 9-10 yrs. In continuous phenotypic features, we used subscale or total scores where available. For example, subscale scores exemplifying different types of sleep-related disorders from the Munich Chronotype Questionnaire. Metrics directly quantifying mental health symptoms were excluded since we aimed to predict cases of mental illness without using symptoms as the latter tend to inflate predictive performance and narrow the utility of findings. The feature set was then partitioned into training and test sets conforming at the participant level with case/control partitions described above (**Construction of participant samples for cases with externalizing disorders**; **Figure 1**). Pre-processing of features was then performed separately in the training and test sets to minimize bias. First, features with >35% missing values were discarded, where prior research shows that good results may be obtained with ML methods with imputation up to 50% missing data. (37) Nominal or ordinal variables were one-hot encoded to transform them into discrete variables. Continuous variables were trimmed to [mean +/- 3] standard deviations to remove outliers and all features scaled in the interval [0,1] with MinMaxScaler. Missing values were imputed using non-negative matrix factorization (NNMF), a mathematically-proven imputation method that minimizes the cost function of missing data rather than assuming zero values. It captures both global and local structure in the data effectively and is particularly suitable for large-scale multimodal data having been demonstrated to perform well regardless of the underlying pattern of missingness. (38, 39, 40) **Supplementary Table 2** shows the number and percentage of observations in each variable which were trimmed and filled with NNMF for the training and test sets, respectively. After imputation with NNMF, phenotypic variables lacking summary scores were reduced to a summary metric or index using feature agglomeration to produce a final set of (*n=*763) non-neural metrics. As described above, neural metrics (*n*=5,014) had already been processed and underwent quality control by the ABCD study team and were therefore not pre-processed with the exception of scaling with the MinMaxScaler, again performed separately in the training and test partitions. There were no missing neural features. The final combined, multimodal feature set including all feature types contained 5,777 features.

### Overview of predictive analytic pipeline

We used deep learning with artificial neural networks to predict cases of ADHD, ODD and CD at 11-12 yrs. In total, we performed 12 experiments, predicting new onset and all prevailing cases for each of the three disorders using a) all available multimodal features and b) only neural features. Deep learning models were implemented with *k*-fold cross-validation and trained by an AI algorithm that jointly performed feature selection and optimized across the hyperparameters in an automated manner. Typically, ∼40,000 model fits were performed during training in each experiment. Model training was terminated based on the Bayes Information Criterion (BIC), an information theoretic metric. After training, final models obtain from the optimized training process were tested for their ability to generalize in the holdout, unseen test set and performance statistics of AUROC, accuracy, precision and recall, and ROC curves computed and reported for these final, optimized models. We also computed and report the relative importance of final predictors to making case predictions using the Shapley Additive Explanations (SHAP) technique. Detailed explanations of these methods are provided below. Code for predictive analytics may be accessed at the de Lacy Laboratory GitHub: https://github.com/delacylab/integrated_evolutionary_learning

### Coarse feature selection

We performed coarse feature selection individually for each of the six experimental samples prior to beginning model training to reduce the number of features entering the deep learning pipeline in a principled, optimized manner. This identified subsets of the total 5,777 features with non-zero relationships with the predictive target. First, a simple filtering process was performed in which χ^2^ (categorical features) and ANOVA (continuous features) statistics and mutual information metric (all features) were computed to quantify the relationship between all features and the target, where the target (ADHD, ODD, CD) was represented by a categorical vector in [0,1]. Any feature with a non-zero relationship (either positive or negative) with the target was retained. Further feature selection was then performed on these filtered feature subsets using the Least Absolute Shrinkage and Selection Operator (LASSO) algorithm. This popular regularization technique based in linear regression efficiently selects a reduced set of features by forcing certain regression coefficients to zero. The LASSO algorithm has a hyperparameter (commonly called the α) that governs the degree of penalization (shrinkage) that will be imposed on the features and thereby influences results. In order to optimize across this hyperparameter, we implemented the LASSO with our AI meta-learning algorithm Integrated Evolutionary Learning (IEL) to tune the α hyperparameter in the same manner as described below in **Integrated Evolutionary Learning for deep learning optimization**.

The number of features retained for each of the six experimental samples after each step described above for the coarse feature selection process may be seen in **Table 3**. Specific features selected in the optimized LASSO regularization and the resulting univariate coefficients between each of these features and the target vectors (ADHD, ODD, CD) for each participant sample (new onset and all prevailing cases at 11-12 yrs) may be viewed in **Supplementary Table 3a-f**. Each feature set selected by the LASSO then entered the deep learning pipeline.

**Table 3:** Feature sets after coarse feature selection for each experiment. The total set of 5,777 multimodal input features was reduced via coarse feature selection in a two-step process of filtering followed by optimized regularization with the LASSO algorithm. This table displays the number of remaining features after filtering and regularization for each target (ADHD, ODD, CD) and participant sample type (new onset cases and all prevailing cases at age 11-12 years). Detailed tables showing the univariate coefficients between each feature selected by LASSO-based regularization and each target may be viewed in **Supplementary Table 3a-f**.

|  | Number of features after filtering | Number of features after selection with LASSO |
| --- | --- | --- |
| ADHD, new onset 11-12 yrs | 4,272 | 54 |
| ODD, new onset 11-12 yrs | 4,314 | 76 |
| CD, new onset 11-12 yrs | 4,303 | 77 |
| ADHD, all prevailing cases 11-12 yrs | 4,388 | 63 |
| ODD, all prevailing cases 11-12 yrs | 4,401 | 81 |
| CD, all prevailing cases 11-12 yrs | 4,460 | 59 |

### Deep learning with artificial neural networks

We used deep learning to predict cases of ADHD, ODD and CD in each type of participant sample (new onset and all prevailing cases at 11-12 yrs). To predict only future cases of externalizing disorders, candidate predictors collected at 9-10 yrs were solely used to predict cases at 11-12 yrs (**Figure 2**). We further recapitulated each experiment after restricting the set of candidate predictors to 5,014 neural features to construct neural-only models to compare their performance to that obtained with multimodal features.

**Figure 2:**
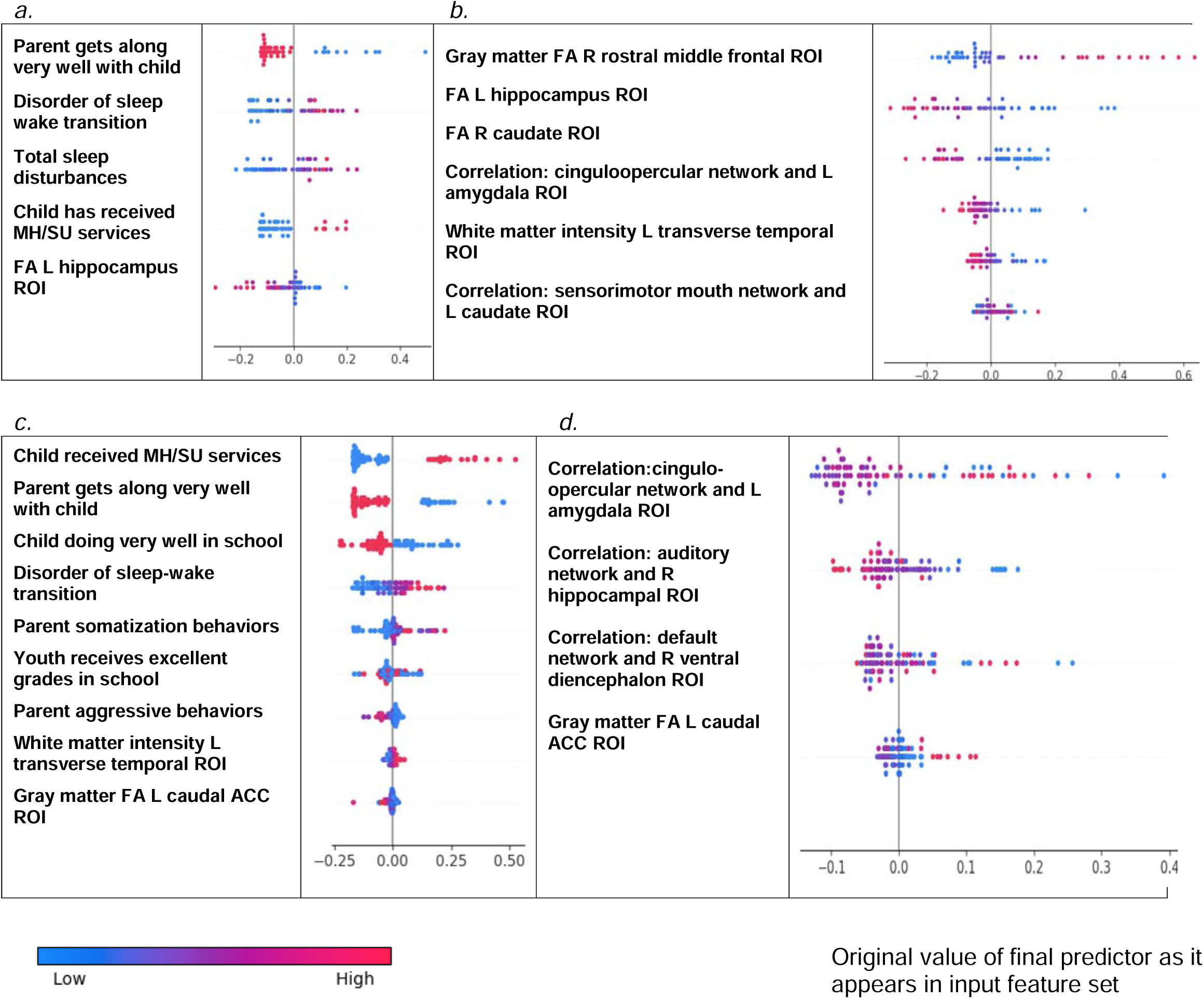
Analytic schema. Features assessed at 9-10 yrs were used to predict new onset and all prevailing cases of ADHD, ODD and CD present at 11-12 yrs.

In each case, we trained artificial neural networks using the AdamW algorithm with 3 layers, 300 neurons per layer, early stopping (patience = 3, metric = validation loss) and the Relu activation function. The last output layer contained a conventional softmax function. Learning hyperparameters (**Table 3**) were tuned with IEL as detailed below. Deep learning models were encoded with TensorFlow embedded in custom Python code.

### Integrated Evolutionary Learning for optimization across hyperparameters and fine feature selection

ML algorithms typically have hyperparameters that control learning, where their settings can strongly affect performance. In many approaches, these hyperparameters are used at their default settings or manually tuned using ‘rules of thumb’ and a restricted number of model fits are explored, introducing the possibility of bias and potentially limiting the solution space. (41, 42, 43) To address this issue, we previously developed an AI technique called Integrated Evolutionary Learning (IEL) which can improve the performance of ML predictive algorithms in tabular data by up to 20-25% versus the use of default model hyperparameters. (44) IEL is a form of computational intelligence or metaheuristic based on an evolutionary algorithm that instantiates the concepts of biological evolutionary selection in computer code. It optimizes across the hyperparameters of the deep learning algorithm by adaptively breeding models over hundreds of learning generations by selecting for improvements in a fitness function (here, the Bayes Information Criterion, BIC).

For each experiment, the deep learning algorithm was nested inside IEL, which initialized the first generation of 100 models with randomized hyperparameter values or ‘chromosomes’. Hyperparameter settings (**Table 4**) were subsequently recombined, mutated or eliminated over successive generations. In recombination, ‘parent’ hyperparameters were arithmetically averaged to form ‘children’. In mutation, settings were shifted with the range of possible values shown in **Table 4**. After the first training generation, the BIC was computed for each of the 100 solutions. The 60 best models (highest BIC) were identified and 40 of these recombined by averaging the hyperparameter setting after a pivot point at the midpoint to produce 20 ‘child’ models. The remaining 20 were mutated to produce the same number of child models by shifting the requisite hyperparameter by the mutation shift value (**Table 4**). The remaining 40 models were discarded. The next generation of models was then formed by adding 60 new models with randomized settings and adding these to the 40 child models retained from the initial generation. Thereafter, IEL continued to recombine, mutate and discard 100 models per generation in a similar fashion to minimize the BIC until the latter fitness function plateaued. With 100 models fitted per generation, IEL typically fits ∼40,000 models per experiment over ∼400 learning generations.

**Table 4:** Hyperparameter settings optimized with Integrated Evolutionary Learning. Optimization across the hyperparameters of learning rate, Beta 1 and Beta 2 was conducted for deep learning with artificial neural networks within the ranges shown.

| Hyperparameters | Range | Mutation Shift |
| --- | --- | --- |
| Learning rate | 0.00001-0.01 | 0.0001 |
| Beta 1 | 0.9-0.999 | 0.001 |
| Beta 2 | 0.9-0.999 | 0.001 |

IEL jointly performs this optimization process across hyperparameter settings with automated feature selection and mitigate the risk of overfitting and identify predictors that perform best. For each experiment, IEL selects from among available candidate predictors after coarse feature selection (**Coarse feature selection, Supplementary Table 3**). A random number of features in the range [2-50] was randomly seeded for each model in the initial learning generation. After computing the fitness function, feature sets from the best-performing 60 models were allocated to child models and other feature sets discarded. As with hyperparameter tuning, this process was repeated for succeeding generations until the BIC plateaued.

IEL implements recursive learning to facilitate computational efficiency. After training until the BIC plateaued, we determine the elbow of the fitness function plotted versus number of features and re-start learning with a warm start. The feature set available after this warm start is constrained to that subset of features, thresholded by their importance, corresponding to the fitness function elbow. Learning then proceeds by thresholding features available for learning at the original warm start feature importance + 2 standard deviations. In addition, the number of models per generation is reduced to 50 and 20 models are recombined and 10 models are mutated. Otherwise, training after the warm start uses the same principles as detailed above.

### Cross validation

Deep learning models were fit within IEL using stratified *k*-fold cross validation i.e. every one of the 100 models in each learning generation within IEL was individually trained and validated using cross-validation. As described above, IEL allows the number of features used to fit each model to differ within each model in every generation in the range [2-50]. Accordingly, *k* (the number of splits) was set as the nearest integer above [sample size/number of features]. Cross validation was implemented with the scikit-learn StratifiedKFold function.

### Testing for generalization in holdout, unseen test data and performance measurement

Final, optimized models generated in the IEL-supervised training process were tested on the held-out, unseen test set for each sample and disorder by applying the requisite hyperparameter settings and selected features to the test set. The area under the receiver operating curve (AUROC), accuracy, precision, and recall were computed for test set models using standard Sci-Kit learn libraries. The most accurate models are presented in **Results**. The threshold for prediction probability was 0.5 and receiver operating characteristic (ROC) curves are also provided for each experiment (**Supplementary Figures 1** and **2**).

### Feature importance determination

Shapley Additive Explanations (SHAP) values were computed using the SHAP toolbox (https://shap.readthedocs.io/en/latest/) to determine the relative importance of each feature to predicting cases in each experiment for ADHD, ODD and CD. SHAP is a game theoretic approach commonly used in ML to explain the output of any ML model including ‘black box’ estimators such as artificial neural networks and is resistant to multicollinearity. (45) It unifies prior methods such as LIME, Shapley sampling values and Tree Interpreter.

## RESULTS

### Overview

All study results detailed below are from testing the final model obtained after IEL optimization for generalization in a holdout, unseen test dataset for each experiment. We present parallel sets of results for each disorder (ADHD, ODD, CD) in predicting new onset cases at 11-12 yrs and all prevailing cases at 11-12 yrs. Only features collected at 9-10 yrs are input to deep learning to make predictions. Therefore, all results represent predictions of future case status. For each disorder, results are presented for standard ML performance metrics and quantification of feature importance for a) <u>multimodal</u> models constructed using all types of input features; and b) <u>neural-only</u> models as follows:

- Performance statistics: accuracy, precision, recall and the AUROC. ROC curves may be viewed in Supplementary Figures 1 and 2.
- Final predictors ranked in order of importance by their group-level SHAP score (average absolute value across the participant sample) and the mean predictor importance (group-level SHAP score) for the requisite experiment.
- Summary SHAP plots that graph individual-level final predictor importance (SHAP scores) for each member of the participant sample. SHAP summary plots are also used to determine the directionality of the relationship between the predictor and case status.

### ADHD

Using multimodal data obtained at 9-10 yrs, deep learning optimized with IEL predicted future, new onset cases of ADHD at 11-12 yrs with ∼86% accuracy, 0.92 AUROC and precision and recall >80%. When predicting all prevailing cases at 11-12 yrs, performance improved to ∼94% accuracy, ∼0.99 AUROC and precision and recall >90%. When only neural features were used, performance fell by ∼6-9% in predicting new onset cases and up to 40% in prevailing cases. Neural-only models predicted new onset cases moderately well with 79% accuracy, 0.841 AUROC and precision and recall ∼74%. Performance in predicting prevailing cases with neural-only features was poor, with ∼64% accuracy, 0.654 AUROC and <60% precision and recall.

The presence of a disorder of excessive somnolence was the most important predictor of new onset case status in ADHD with parent-child conflict present to a lesser degree. The model that predicted all prevailing cases at 11-12 yrs was more complex. The most important predictors were whether the child had received mental health or substance abuse services prior to assessment at 9-10 yrs and the total level of parental behavioral problems. This was followed by conflict between parent and child, presence of a disorder of sleep-wake transition or excessive somnolence, and the level of parental externalizing behaviors. For both new onset and all prevailing cases, how well the child functioned at school and specifically having excellent grades in school had an inverse predictive relationship with ADHD case status.

**Table 5:**
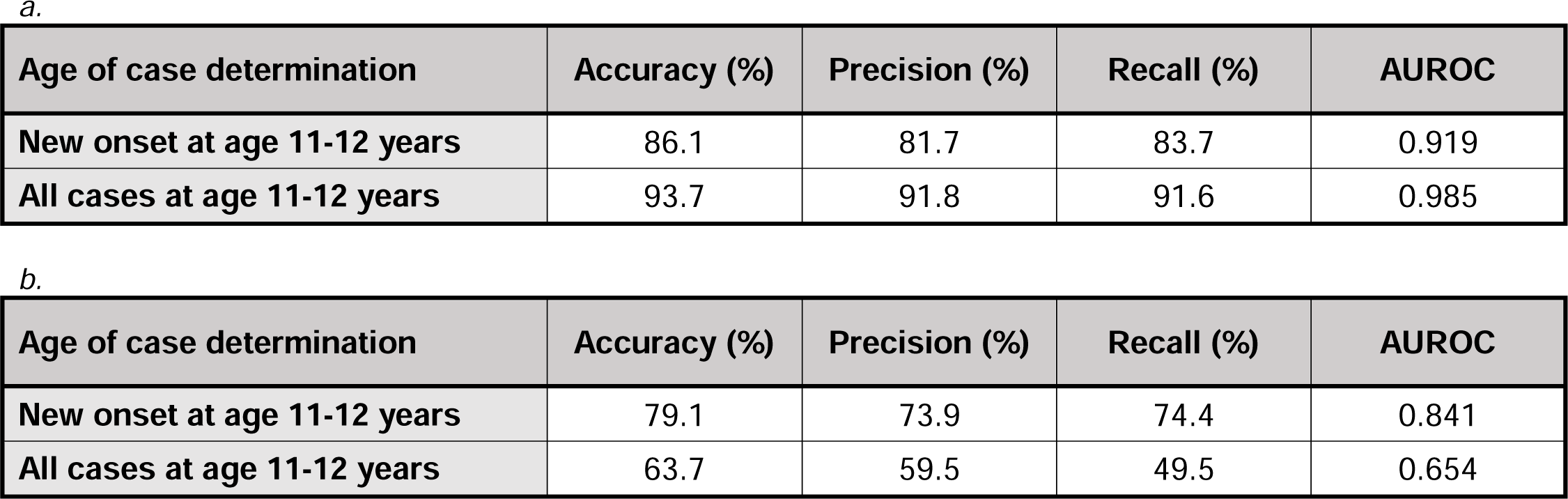
Performance of deep learning optimized with Integrated Evolutionary Learning in predicting cases of ADHD using multimodal and neural-only feature types. Performance statistics of accuracy, precision, recall and the AUROC are shown for the most accurate model obtained with deep learning optimized with Integrated Evolutionary Learning using a) multimodal features and b) only neural features. We used features obtained at 9-10 years of age to predict new onset cases of ADHD at 11-12 years of age as well as all prevailing cases at 11-12 years of age. Corresponding ROC curves may be viewed in **Supplementary Figures 1** and **2**.

| Age of case determination | Accuracy (%) | Precision (%) | Recall (%) | AUROC |
| --- | --- | --- | --- | --- |
| New onset at age 11-12 years | 86.1 | 81.7 | 83.7 | 0.919 |
| All cases at age 11-12 years | 93.7 | 91.8 | 91.6 | 0.985 |

**Table 5: Performance of deep learning optimized with Integrated Evolutionary Learning in predicting cases of ADHD using multimodal and neural-only feature types**
| Age of case determination | Accuracy (%) | Precision (%) | Recall (%) | AUROC |
| --- | --- | --- | --- | --- |
| New onset at age 11-12 years | 79.1 | 73.9 | 74.4 | 0.841 |
| All cases at age 11-12 years | 63.7 | 59.5 | 49.5 | 0.654 |

In prevailing cases, this was joined by the child’s level of prosocial behaviors. In multimodal models where all feature types were available, the optimization process run by IEL preferentially selected psychosocial features with no cognitive, neural or biological metrics present in final, optimized models. Group-level importances for multimodal model predictors (averaged across the participant sample) were in the range [0.009, 0.20] and the mean importance for each experiment in the range [0.06, 0.12].

**Table 6:** Final predictors of cases of ADHD at age 11-12 years. Final predictors of cases of all prevailing cases of ADHD at 11-12 years as well as new onset cases only at 11-12 years of age are shown for the most accurate models obtained using deep learning optimized with IEL obtained with a) multimodal features and b) only neural features. Final predictors are ranked in order of importance where the relative importance of each predictor is computed with the Shapley Additive Explanations technique and presented here averaged across all participants in the sample. Features in red indicate an inverse relationship with ADHD verified with the Shapley method. MH = mental health; SU=substance use; SST = Standard Stop Signal task; MID = Monetary Incentive Delay task; ROI = region of interest; FA = fractional anisotropy; WM = white matter; GM = gray matter.

| Age of case determination | Ranked Final Predictors | Importance |
| --- | --- | --- |
| New onset at age 11-12 years | Disorder of excessive somnolence | 0.2038 |
|  | Does very well at school | 0.1423 |
|  | Parent reports some conflict with child | 0.0796 |
|  | Youth gets excellent grades at school | 0.0567 |
|  | Mean | 0.1206 |
| All cases at age 11-12 years | Child has received MH/SU services in last 6 months | 0.1638 |
|  | Parent total behavioral problems | 0.1061 |
|  | Parent reports some conflict with child | 0.0725 |
|  | Disorder of sleep-wake transition | 0.0665 |
|  | Youth performs very well in school | 0.0617 |
|  | Disorder of excessive somnolence | 0.0375 |
|  | Youth receives excellent grades in school | 0.0234 |
|  | Prosocial behaviors mean score | 0.0147 |
|  | Parent externalizing problems | 0.0091 |
|  | Mean | 0.0617 |

**Table 6: Final predictors of cases of ADHD at age 11-12 years**
| Neural data type | Ranked Final Predictors | Importance |
| --- | --- | --- |
| New onset at age 11-12 years | T1 intensity in brain stem ROI | 0.0890 |
|  | Correlation between ventral attention network and right ventral diencephalon ROI | 0.0532 |
|  | SST any stop vs correct go contrast in left pars opercularis ROI | 0.0411 |
|  | SST incorrect stop vs correct go contrast in left lingual ROI | 0.0364 |
|  | T1 intensity WM for left lateral occipital ROI | 0.0309 |
|  | Cortical thickness in mm of right transverse temporal ROI | 0.0304 |
|  | MID loss anticipation vs neutral contrast in right supramarginal ROI | 0.0271 |
|  | Average FA in GM right caudal ACC ROI | 0.0147 |
|  | Mean | 0.0400 |
| All cases at age 11-12 years | GM FA in right caudal middle frontal ROI | 0.0446 |
|  | Cortical area in mm<sup>2</sup> of left inferior parietal ROI | 0.0259 |
|  | MID small loss vs neutral contrast in right inferior temporal ROI | 0.0192 |
|  | SST incorrect go vs incorrect stop contrast in left lingual ROI | 0.0134 |
|  | Cortical thickness in mm of left pars triangularis ROI | 0.0129 |
|  | SST incorrect stop vs correct go in left lingual ROI | 0.0046 |
|  | Mean | 0.0200 |

In interpreting the neural-only experiments, we observed little overlap between the final, optimized models for new onset and all prevailing cases of ADHD. The only common feature was a negative relationship between case status and SST contrast in the left lingual ROI, though the contrast effect differed between incorrect stop vs correct go (new onset) and incorrect go vs incorrect stop (prevailing cases). In new onset cases, the most prominent positive predictor was the correlation between the ventral attention network and right ventral diencephalon ROI, followed by SST contrast in the left pars opercularis and cortical thickness in the right transverse temporal ROI. Structural differences in the brain stem, left lateral occipital white matter and right caudal ACC along with MID contrast in the right supramarginal ROI were negative predictors of new onset case status. The neural-only model of all prevailing ADHD cases was less reliable, with an AUROC of 0.654, but we found structural features in the right caudal middle frontal and left pars triangularis ROIs predicted case status with inverse relationships with cortical area of the left parietal ROI and MID loss contrast in the right inferior temporal ROI. Group-level importances for neural-only model predictors were in the range [0.02,0.04] and the mean importance for each experiment in the range [0.0046, 0.089], both representing lower importance ranges than multimodal models.

We further computed and plotted individual-level SHAP values to quantify the dispersion of importances across individuals and assess the directionality of the relationship between final predictors and clinical case status (**Figure 3**). In these summary plots, each data point represents an individual participant and the colorization reflects the original value of the predictor as an input feature. Thus, discrete-valued features appear as red or blue, whereas a continuous feature appears as a color gradient from low to high.

**Figure 3:**
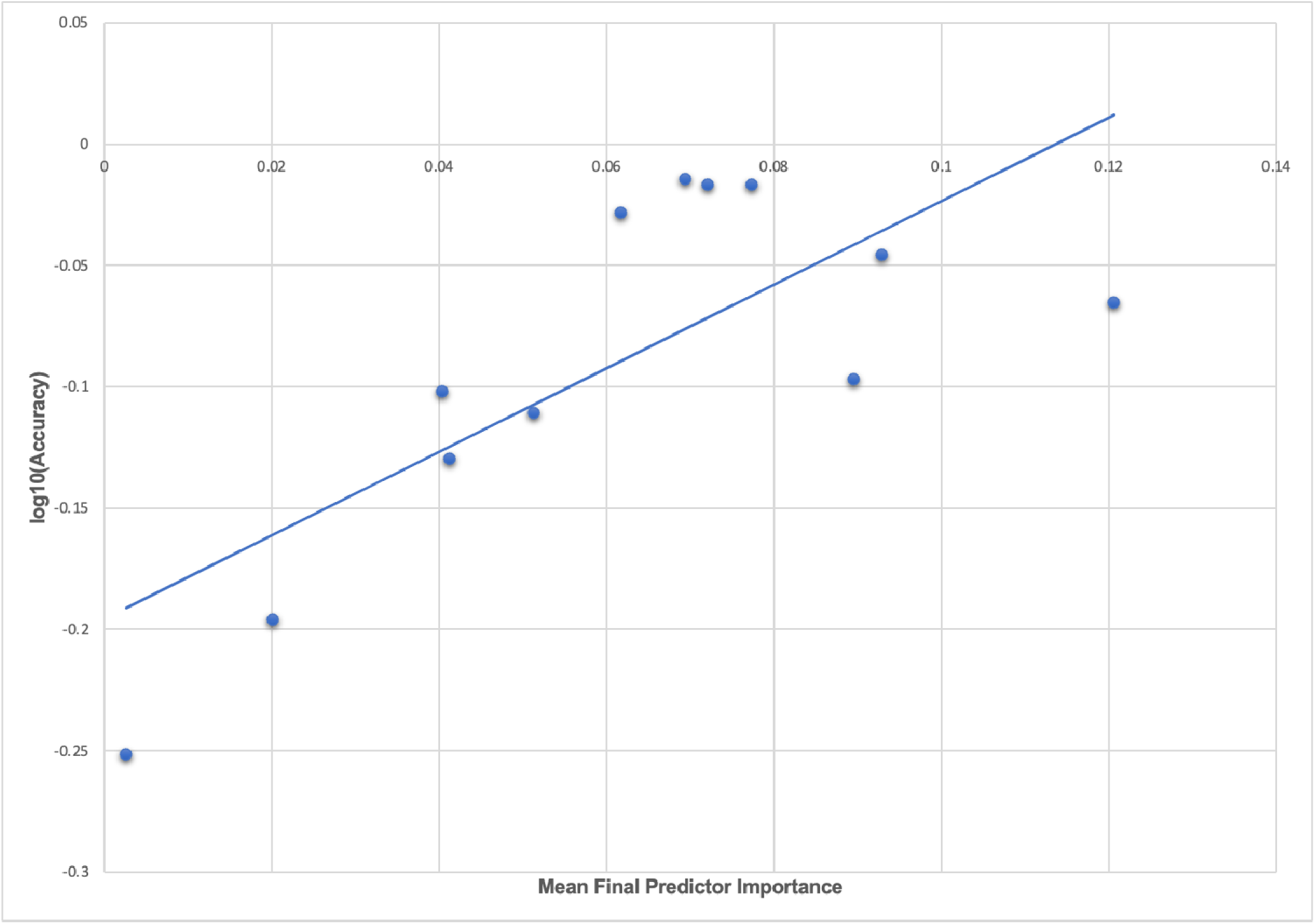
Individual-level importances of final predictors of ADHD in early adolescence. Summary plots are presented of the importance of each final predictor (computed with the Shapley Additive Explanations technique) on an individual subject level to predicting ADHD with new onset at 11-12 yrs with a) multimodal features and b) only neural features and in all prevailing cases of ADHD at 11-12 yrs with c) multimodal features and d) only neural features. The color gradient represents the original value of each feature (metric) where red = high and blue = low. Discrete (binary) features appear as red or blue, while continuous features appear as a color gradient.

Individual-level importances in multimodal predictive models of both new onset and prevailing cases of ADHD were typically more widely dispersed than in neural-only models. Further, wider dispersions across the participant samples were observed for the more important predictors.

### Oppositional Defiant Disorder

In ODD, predictive models performed strongly using multimodal features. In new onset cases, we achieved accuracy of ∼97%, AUROC of 0.996 and precision and recall ≥94% with 96% accuracy, AUROC of 0.988 and precision and recall ≥95% when predicting all prevailing cases at 11-12 yrs. In neural-only models we observed similar phenomena as in ADHD: performance fell substantially with relatively better performance in predicting new onset vs prevailing cases. When only neural features were used, performance fell by ∼20% in predicting new onset cases and up to ∼40% in prevailing cases. Neural-only models predicted new onset cases moderately well with 74% accuracy, 0.792 AUROC and precision and recall ≥65%. Performance in predicting prevailing cases with neural features was poor, with ∼56% accuracy, 0.567 AUROC and <55% precision and recall.

**Table 7:** Performance of deep learning optimized with Integrated Evolutionary Learning in predicting cases of ODD using multimodal and neural-only feature types. Performance statistics of accuracy, precision, recall and the AUROC are shown for the most accurate model obtained with deep learning optimized with Integrated Evolutionary Learning using a) multimodal features and b) only neural features. We used features obtained at 9-10 years of age to predict new onset cases of ODD at 11-12 years of age as well as all prevailing cases at 11-12 years of age. Corresponding ROC curves may be viewed in **Supplementary Figures 1** and **2**.

| Age of case determination | Accuracy (%) | Precision (%) | Recall (%) | AUROC |
| --- | --- | --- | --- | --- |
| New onset at age 11-12 years | 96.8 | 96.8 | 93.5 | 0.996 |
| All cases at age 11-12 years | 96.2 | 94.8 | 95.5 | 0.988 |

**Table 7: Performance of deep learning optimized with Integrated Evolutionary Learning in predicting cases of ODD using multimodal and neural-only feature types**
| Age of case determination | Accuracy (%) | Precision (%) | Recall (%) | AUROC |
| --- | --- | --- | --- | --- |
| New onset at age 11-12 years | 74.2 | 69.4 | 64.5 | 0.792 |
| All cases at age 11-12 years | 56.1 | 53.9 | 27.2 | 0.567 |

Whether the youth had ever received mental health or substance abuse services prior to assessment at age 9-10 yrs was the most important predictor of new onset case status in ODD followed by the presence of a disorder of excessive somnolence or sleep-wake transition. Additional important predictors were parental factors: the presence of nerves or a nervous breakdown problem and levels of externalizing or aggressive behaviors. Youth prosocial behaviors exhibited an inverse relationship with case status. Features that predicted all prevailing cases at 11-12 yrs included a number of final predictors that were the same or thematically similar: whether the child had received mental health or substance abuse services in the last 6 months (the most important predictor), total sleep disturbances, disorder of sleep-wake transition, parent externalizing behaviors and an inverse relationship with prosocial behaviors. Final predictors that differed in this model were the youth’s mother having a depression problem and whether either parent had sought treatment for a mental or emotional problem. Of note, the latter predictor had an inverse relationship with case status, suggesting it was related to (an) untreated mental problem(s). In multimodal models where all feature types were available, the optimization process run by IEL preferentially selected psychosocial features with no cognitive, neural or biological metrics present in final, optimized models. Group-level importances for multimodal model predictors (averaged across the participant sample) were in the range [0.003, 0.18] and the mean importance for each experiment at ∼0.07.

In neural-only models, the future onset of ODD at 11-12 yrs was predicted by a model (with moderately strong performance at AUROC = 0.79) containing only rsfMRI-derived correlations. Strikingly, every final predictor represented a correlation metric between a cortical network and subcortical ROI, emphasizing networks involved in salience, executive function, spatial memory and task performance. Of note, all neural features with positive relationships with the onset of ODD were in the left hemisphere and those with inverse relationships with case status in the right hemisphere. As noted above, the neural-only model predicting all prevailing cases of ODD at 11-12 yrs exhibited poor performance (AUROC = ∼0.567) and as such it cannot be considered reliable. It consisted of two structural gray matter features: fractional anisotropy of the right lateral orbitofrontal ROI and cortical area of the left inferior parietal ROI. Group-level importances for neural-only model predictors (averaged across the participant sample) were in the range [0.0007, 0.075] and the mean importance for each experiment in the range [0.0026, 0.0410].

**Figure 4:**
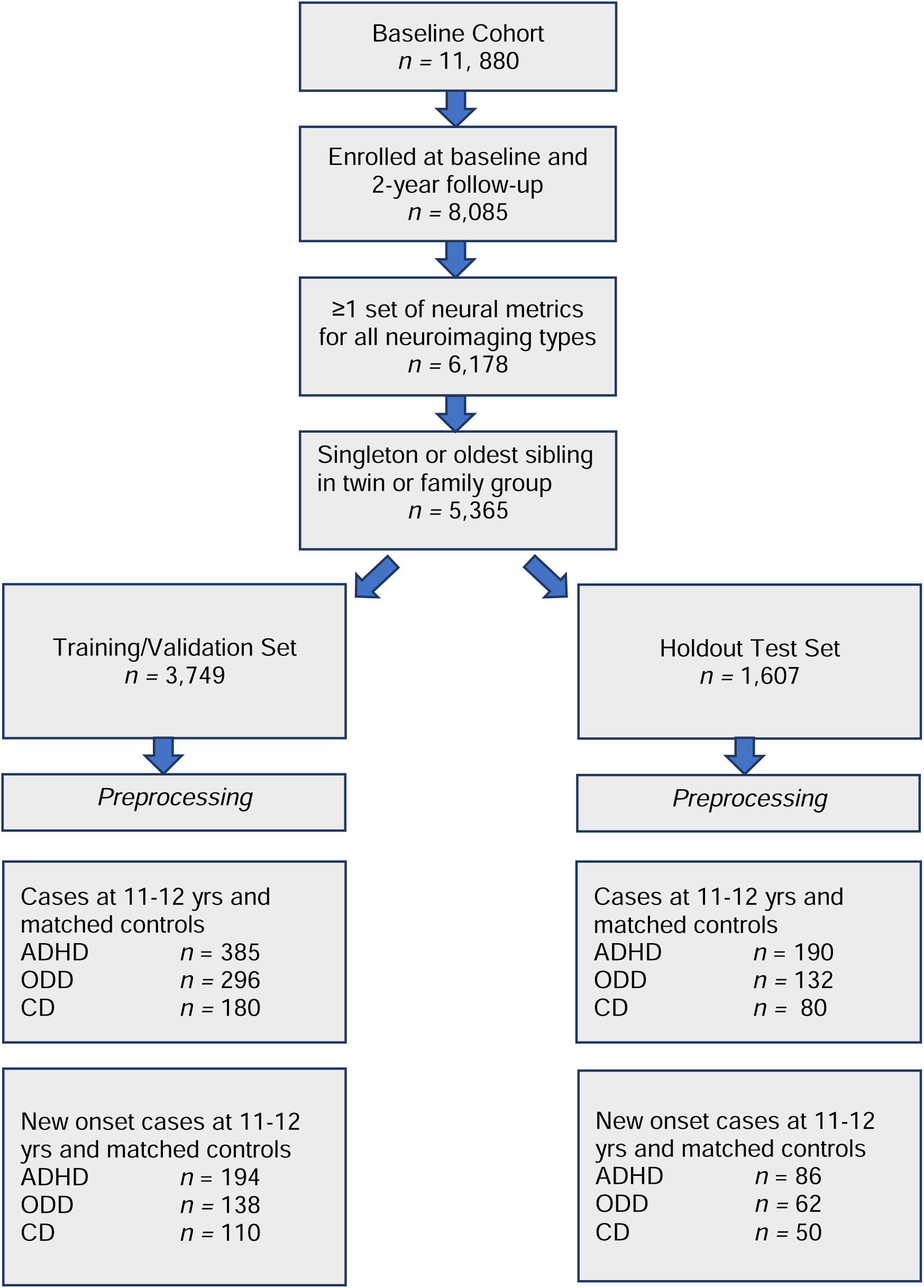
Individual-level importances of final predictors of Oppositional Defiant Disorder in early adolescence. Summary plots are presented of the importance of each final predictor (computed with the Shapley Additive Explanations technique) on an individual subject level to predicting ODD with new onset at 11-12 yrs with a) multimodal features and b) only neural features and in all prevailing cases of ODD at 11-12 yrs with c) multimodal features and d) only neural features. The color gradient represents the original value of each feature (metric) where red = high and blue = low. Discrete (binary) features appear as red or blue, while continuous features appear as a color gradient.

**Table 8:**
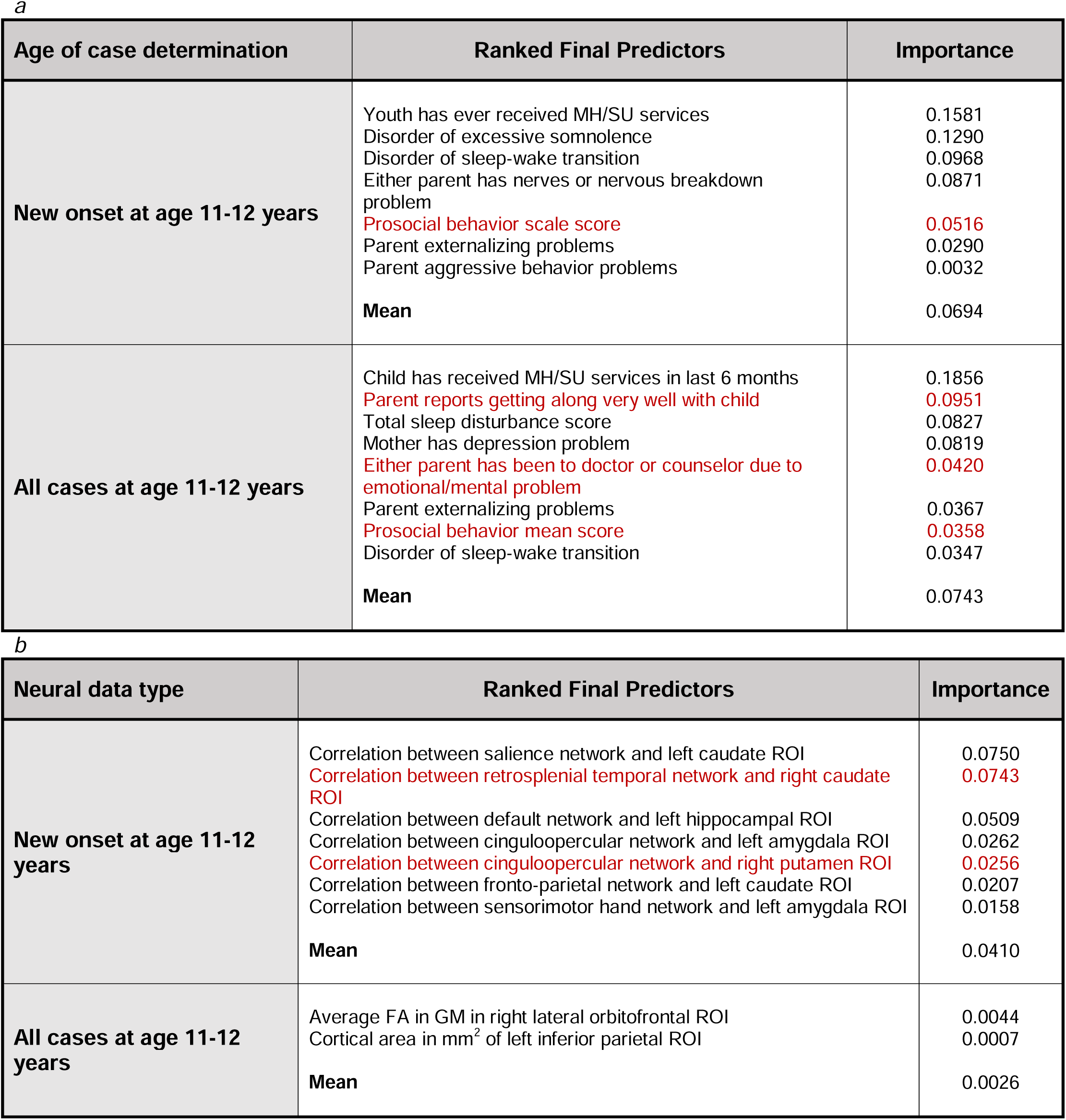
Final predictors of cases of ODD at age 11-12 years. Final predictors of cases of all prevailing cases of ODD at 11-12 years as well as new onset cases only at 11-12 years of age are shown for the most accurate models obtained using deep learning optimized with IEL obtained with a) multimodal features and b) only neural features. Final predictors are ranked in order of importance where the relative importance of each predictor is computed with the Shapley Additive Explanations technique and presented here averaged across all participants in the sample. Features in red indicate an inverse relationship with ODD verified with the Shapley method. MH = mental health; SU=substance use; ROI = region of interest; GM = gray matter.

As observed in ADHD, individual-level importances in multimodal predictive models of both new onset and prevailing cases of ODD were typically more widely dispersed than in neural-only models. Further, wider dispersions across the participant samples were observed for the more important predictors.

### Conduct Disorder

Deep learning optimized with IEL predicted future, new onset cases of CD at 11-12 yrs with ∼90% accuracy, 0.92 AUROC and precision and recall >85% using multimodal features assessed at 9-10 yrs. In predicting all prevailing cases at 11-12 yrs, performance improved further to ∼96% accuracy, ∼0.99 AUROC and precision and recall ≥95%. This strong predictive performance represented the best overall performance among the three externalizing conditions. When only neural features were used, performance fell by ∼10% in predicting new onset cases and up to 20% in prevailing cases. However, this is in the context of neural-only models achieving moderately strong performance in predicting new onset cases with 80% accuracy, 0.808 AUROC and precision and recall >70%. Performance in predicting prevailing cases with neural-only features was also moderately strong with ∼78% accuracy, 0.816 AUROC and >70% precision and recall.

**Table 9:** Performance of deep learning optimized with Integrated Evolutionary Learning in predicting cases of conduct disorder using multimodal and neural-only feature types. Performance statistics of accuracy, precision, recall and the AUROC are shown for the most accurate model obtained with deep learning optimized with Integrated Evolutionary Learning using a) multimodal features and b) only neural features. We used features obtained at 9-10 years of age to predict new onset cases of CD at 11-12 years of age as well as all prevailing cases at 11-12 years of age. Corresponding ROC curves may be viewed in **Supplementary Figures 1** and **2**.

| Age of case determination | Accuracy (%) | Precision (%) | Recall (%) | AUROC |
| --- | --- | --- | --- | --- |
| New onset at age 11-12 years | 90.0 | 85.4 | 92.0 | 0.922 |
| All cases at age 11-12 years | 96.3 | 95.1 | 95.0 | 0.990 |

**Table 9: Performance of deep learning optimized with Integrated Evolutionary Learning in predicting cases of conduct disorder using multimodal and neural-only feature types** Performance statistics of accuracy, precision, recall and the AUROC are shown for the most accurate model obtained with deep learning optimized with Integrated Evolutionary Learning using a) multimodal features and b) only neural features.
| Age of case determination | Accuracy (%) | Precision (%) | Recall (%) | AUROC |
| --- | --- | --- | --- | --- |
| New onset at age 11-12 years | 80.0 | 75.7 | 72.0 | 0.808 |
| All cases at age 11-12 years | 77.5 | 71.7 | 75.0 | 0.816 |

The interpretation of predictive models for CD was particularly intriguing. Unlike ADHD and ODD, final predictors of both new onset cases and all prevailing cases at 11-12 yrs using multimodal data did include neural features. New onset cases of CD were predicted by psychosocial features also found in ADHD and ODD (tenor of parent-child relationship, sleep disturbances, mental health treatment prior to age 9-10 yrs) but here these psychosocial factors interacted in an inverse relationship with structural disturbance in the left hippocampal ROI. Similarly, final predictors of all prevailing cases of CD at 11-12 yrs comprised psychosocial features common to ADHD and ODD (prior mental health treatment, tenor of parent-child relationship, sleep disturbances, school performance) but these interacted with structural features in the left transverse temporal white matter and left caudal anterior cingulate cortex gray matter (inverse relationship). A further interesting facet of this latter model was that parent somatization traits were a driver of CD where parent aggressive traits had an inverse relationship with case status. Somatization refers to the expression of mental phenomena as physical (somatic) symptoms, seek medical care for them and placement of an undue focus on the distress caused by physical complaints.

In neural-only models, which performed relatively well in CD, prominent predictors of new onset cases were structural features in the right rostral middle frontal ROI, left hippocampus (as also found in the multimodal model) and right caudate. Less important features included the correlation between the cinguloopercular network and left amygdala (also observed in ODD) and the left transverse temporal ROI (also observed in the multimodal model). Final neural-only predictors of prevailing cases of CD were dominated by cortical-subcortical connectivity features comprising the cinguloopercular network with the left amygdala (also important to new onset prediction), auditory network with right hippocampus and default mode network with right ventral diencephalon. This model was rounded out with structural gray matter differences in the left caudal ACC, also observed in the multimodal model. In both new onset and prevailing cases, there was an emphasis on subcortical structural features and connectivity between cortical networks and subcortical ROIs.

**Figure 5:**
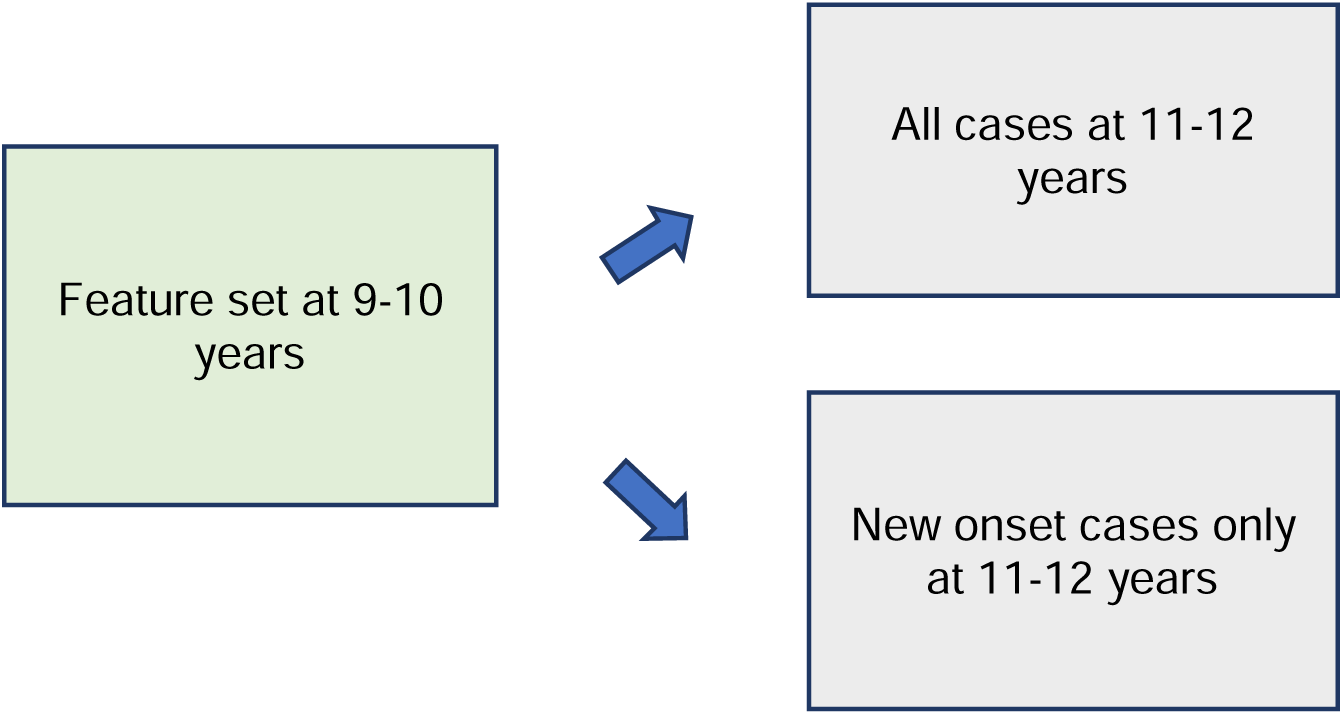
Individual-level importances of final predictors of Conduct Disorder in early adolescence. Summary plots are presented of the importance of each final predictor (computed with the Shapley Additive Explanations technique) on an individual subject level to predicting CD with new onset at 11-12 yrs with a) multimodal features and b) only neural features and in all prevailing cases of CD at 11-12 yrs with c) multimodal features and d) only neural features. The color gradient represents the original value of each feature (metric) where red = high and blue = low. Discrete (binary) features appear as red or blue, while continuous features appear as a color gradient.

**Table 10:** Final predictors of cases of conduct disorder at age 11-12 years. Final predictors of cases of all prevailing cases of CD at 11-12 years as well as new onset cases only at 11-12 years of age are shown for the most accurate models obtained using deep learning optimized with IEL obtained with a) multimodal features and b) only neural features. Final predictors are ranked in order of importance where the relative importance of each predictor is computed with the Shapley Additive Explanations technique and presented here averaged across all participants in the sample. Features in red indicate an inverse relationship with CD verified with the Shapley method. MH = mental health; SU=substance use; ROI = region of interest; FA = fractional anisotropy; WM = white matter; GM = gray matter.

| Age of case determination | Ranked Final Predictors | Importance |
| --- | --- | --- |
| New onset at age 11-12 years | Parent reports gets along very well with child | 0.1126 |
|  | Disorder of sleep wake transition | 0.0996 |
|  | Total sleep disturbances | 0.0949 |
|  | Child has ever received MH/SU services | 0.0927 |
|  | Average FA left hippocampus ROI | 0.0642 |
|  | Mean | 0.0928 |
| All cases at age 11-12 years | Child has received MH/SU services in last 6 months | 0.1593 |
|  | Parent reports getting along very well with child | 0.1442 |
|  | Child is doing very well in school | 0.0958 |
|  | Disorder of sleep-wake transition | 0.0806 |
|  | Parent somatization behaviors score | 0.0455 |
|  | Youth receives excellent grades in school | 0.0408 |
|  | Parent aggressive behavior score | 0.0235 |
|  | T1 intensity WM left transverse temporal ROI | 0.0130 |
|  | Average FA GM left caudal ACC ROI | 0.0123 |
|  | Mean | 0.0683 |

Table 10: Final predictors of cases of conduct disorder at age 11-12 years
| Neural data type | Ranked Final Predictors | Importance |
| --- | --- | --- |
| New onset at age 11-12 years | Average FA GM right rostral middle frontal ROI | 0.1628 |
|  | Average FA left hippocampus ROI | 0.1348 |
|  | Average FA right caudate ROI | 0.1043 |
|  | Correlation between cinguloopercular network and left amygdala ROI | 0.0531 |
|  | T1 intensity WM left transverse temporal ROI | 0.0490 |
|  | Correlation between sensorimotor mouth network and left caudal ROI | 0.0333 |
|  | Mean | 0.0890 |
| All cases at age 11-12 years | Correlation between cinguloopercular network and left amygdala ROI | 0.0960 |
|  | Correlation between auditory network and right hippocampal ROI | 0.0456 |
|  | Correlation between default network and right ventral diencephalon ROI | 0.0444 |
|  | Average FA GM in left caudal ACC ROI | 0.0192 |
|  | Mean | 0.0513 |

As observed in both ADHD and ODD, individual-level importances in multimodal predictive models of both new onset and prevailing cases of CD were typically more widely dispersed than in neural-only models. Further, wider dispersions across the participant samples were observed for the more important predictors.

### The relationship between accuracy and final predictor importance

We computed the mean predictor importance for each experiment to explore the relationship between model accuracy in testing in held-out, unseen data and final predictor importance after optimized, automated feature selection. For example, the average importance of final predictors of new onset ADHD at 11-12 years (**Table 6**). This data may be inspected in **Supplementary Table 4**. Further, we computed the correlation and R^2^ of the relationship between accuracy and mean predictor importance for each experiment described in the present study. Across all experiments, the correlation between accuracy and predictor importance in final, optimized models tested in held-out, unseen data was 72.7% and the R^2^ was 52.8%. This is summarized in **Figure 6** where mean final predictor importance is shown plotted against log(accuracy) to improve scale interpretation, though we note that the reported correlation and R^2^ was computed with accuracy.

**Figure 6:**
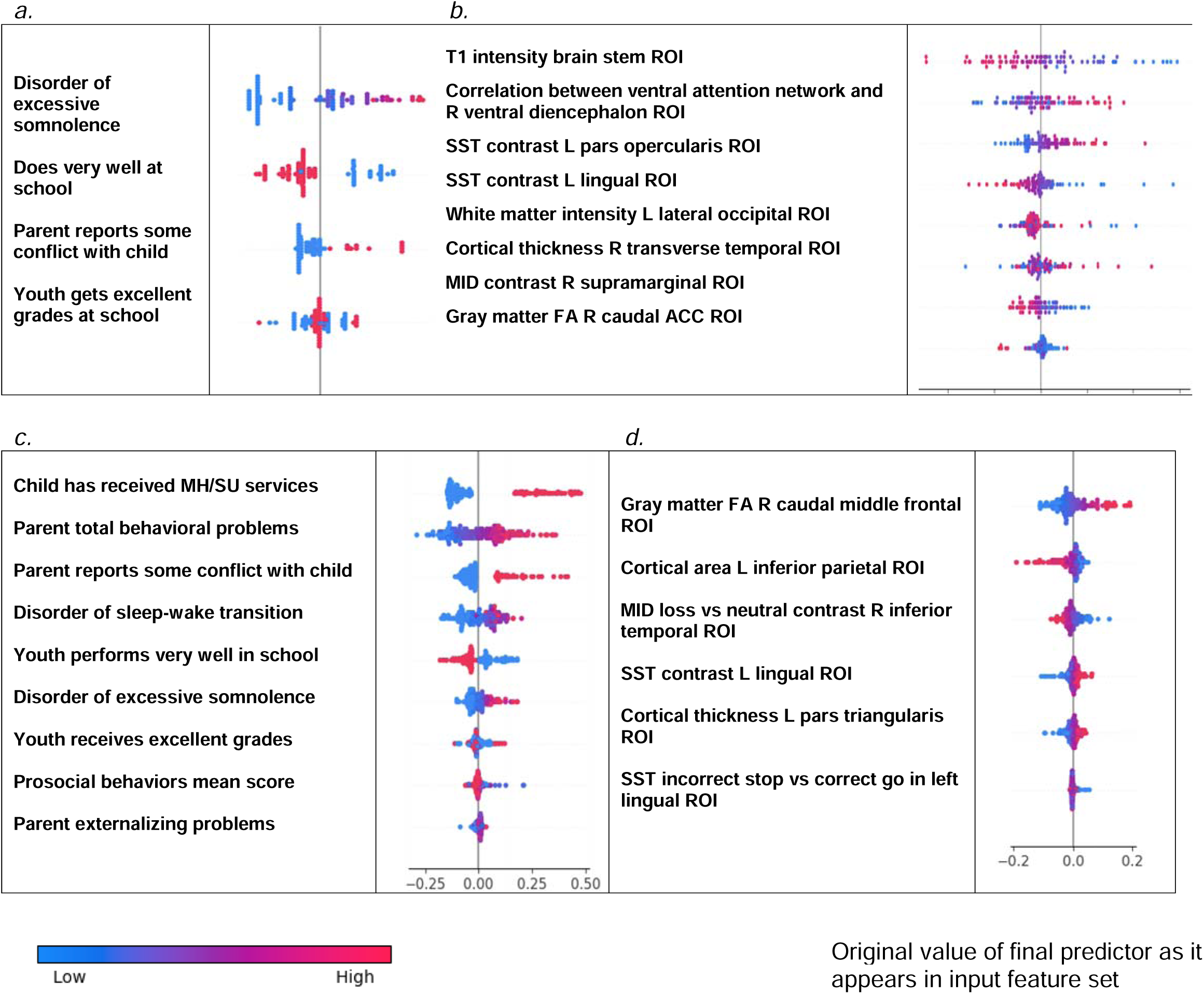
The relationship between accuracy and final predictor importance. Average variable importance computed with the Shapley Additive Explanations technique is shown plotted against the log of prediction accuracy in testing in held-out data for each experiment in the study. The line of best fit obtained with a linear regression is also displayed. Underlying data for this chart may be inspected in **Supplementary Table 4.**

## DISCUSSION

### General observations across externalizing disorders

Using an AI-guided feature selection process, we were able to distil ∼6,000 candidate predictors contributed by children 9-10 yrs and their parents into robust, individual-level models predicting the later (11-12 yrs) onset of ADHD, ODD and CD. This extended prior work in ML prediction of externalizing disorders in adolescence by assessing ∼30x more candidate predictors spanning a wider variety of knowledge domains (cognitive, psychosocial, biological, multiple neural types). By imposing a common pre-processing and analytic design across all three major externalizing disorders in the same participant cohort we were able to directly compare results, quantify the relative predictive performance of multimodal vs neural features and examine the relationship between predictor importance and model accuracy across multiple experiments. To our knowledge, this is the first study using ML to predict the onset of all three major adolescent externalizing disorders and include many types of neural predictors (rsfMRI connectivity; task fMRI effects; diffusion and structural metrics), analyze >200 multimodal features and quantify the relationship between predictor importance and accuracy.

Comparing across experiments, we found that relative predictive performance varied according to disorder and predictor type (psychosocial vs neural). Overall, deep learning optimized with IEL applied to multimodal features achieved strong performance with ∼86-97% accuracy, 0.919-0.996 AUROC and ∼82-97% precision and recall in testing in held-out, unseen data. With multimodal features, performance was slightly stronger in predicting prevailing over new onset cases in ADHD and CD but equivalent in ODD with the strongest performance overall in ODD, followed by CD and then ADHD. Further targeted experiments specifically assessed the standalone predictive ability of multiple neural feature types derived from MRI. After restricting the candidate predictors to 4,777 neural features, we observed that predictive performance dropped substantially across all three disorders, most prominently when predicting all prevailing cases. The small number of prior ML studies in adolescent externalizing disorders that have directly compared the utility of psychosocial vs neural predictors have obtained similar results and performance differentials. (14) However, we would highlight that neural-only features were for the most part able to predict new onset cases with accuracy and AUROC of ∼80%. While not as strong as with multimodal features, this performance compares favorably with the existing literature using ML and biobehavioral features to predict externalizing disorders in adolescents.

To our knowledge, this is the first study to provide directly comparable predictive models of all three major externalizing disorders. In adolescence, ADHD, ODD and CD frequently co-occur in the population, and in adulthood are increasingly co-morbid with mental health conditions such as the internalizing and personality disorders and substance use. It is therefore challenging to assemble a longitudinal cohort where participants have only ADHD, ODD or CD without any co-morbidities, and we are not aware that such a sample exists in adolescence with sufficient participants to enable rigorous ML analyses. Moreover, allowing naturalistic sample overlap among the externalizing conditions may improve translational relevance in that it reflects the clinical population. Here, we adopted a design where all three disorders are predicted in the same cohort using the same methods to allow head-to-head comparison of final predictors and enable the identification of common vs specific predictors across ADHD, ODD and CD in the same population. We found that each set of final predictors was a unique combination of features and differentiated both a) ADHD, ODD and CD from each other and b) future new onset from all prevailing cases. However, there were cross-cutting themes. In predicting case onset, sleep disorders (excessive somnolence, sleep-wake transition and total disturbances) were common, prominent predictors across ADHD, ODD and CD. Sleep disturbances may affect up to ∼40% of elementary school age children and youth with both internalizing and externalizing disorders are at elevated risk. (49, 50) Sleep disturbances have been shown to “precede, predict and significantly contribute” to behavioral issues in ADHD and worsen disruptive behaviors, ODD and CD in adolescence, though links with sleep latency and duration have been variable (19, 20, 21, 22) Here, our findings add to a growing body of work suggesting sleep disturbances may be important intervention targets in elementary school age youth to reduce the later onset of clinical ADHD, ODD and CD. Moreover, we found that daytime somnolence and sleep-wake transition were emphasized in predicting the externalizing disorders in adolescence and not sleep latency or duration. Other themes were shared by two of the three disorders: conflict between parent and child was shared in ADHD and ODD and in the more behaviorally severe disorders (ODD, CD), youth appeared to have come to clinical attention prior to age 9-10 yrs. In our models for all prevailing cases, shared themes were recent mental health treatment for the youth, sleep disturbances and parental burden of various types of behavioral problems and parent-child conflict. Unsurprisingly, therefore, there are thematically common predictors across all three externalizing disorders that also reflect the extant literature. However, disorder-specific predictors did exist that may aid in disambiguating the onset of these conditions. Most strikingly, CD was marked by the importance of structural brain features that interacted with psychosocial predictors and which appeared in neither ADHD nor ODD in multimodal models. As well, neural-only models achieved their best performance in CD over ADHD or ODD. This highlights a potential role for structural neuroimaging in identifying youth at risk for CD, the most severe and disabling of the three disorders, vs ADHD or ODD. In terms of the latter two conditions, school performance was a prominent predictor of the onset of ADHD vs an emphasis on lower levels of prosocial behaviors and parent mental health issues in ODD.

Recent studies suggest that inflated effect sizes in neuroimaging studies of psychopathology and cognitive traits may be responsible for generalization failure, particularly in group-level association studies and smaller participant samples. (55) While there is no exact equivalent to group-level effect size in the individual-level models provided by deep learning with artificial neural networks, predictor importance in the context of accuracy is conceptually similar. We therefore investigated predictor importance at both the group and individual level and its relationship with model performance in generalization testing, finding a moderately strong relationship (R2 ∼53%) between predictor importance and accuracy. Psychosocial predictors in multimodal models had larger importances and wider inter-individual dispersions than those in neural-only experiments, even after extensive optimization and principled feature selection. Collectively, these results suggest that the smaller importances of neural features and their more restricted inter-individual variability were at least related to their weaker performance in predicting cases. Future work will be required to determine whether these phenomena are seen in other disorders and participant samples or if other types of neural features might perform differently in predicting cases of externalizing disorders.

### Predicting the onset of ADHD in early adolescence

ADHD affects up to 10% of school age children and is characterized by inattention, impulsivity and hyperactivity. It is a developmental disorder which shows markedly increasing prevalence from late elementary school through adolescence and is treatable. Thus, the early detection of children at risk for new onset is of substantial interest. There have been a number of ML multimodal predictive studies in adolescent ADHD, predominantly cross-sectional. National-level cohorts have offered large sample sizes to enable ML but typically a smaller range of psychosocial/demographic candidate predictors. For example, Garcia-Argibay et al analyzed 22 candidate predictors in Swedish registry data (*n*=238,696), achieving moderate performance with deep learning (accuracy: 69%, AUROC: 0.75) and identifying top predictors of having a parent with criminal convictions or relative with ADHD, male sex, number of academic subjects failed and speech/learning disabilities. (23) In a Japanese sample (*n =* 45,779), Maniruzzaman et al identified family structure, insurance age, sex, medical conditions, mental health symptomatology as significant among 19 psychosocial candidate predictors (accuracy: 86%, AUROC: 0.94). (24) Using a British school-based cohort, Ter-Minassian et al were able to access a wider range of 68 candidate predictors and found school attendance, social-emotional development level, writing performance, male sex and problem solving/reasoning to be most important in predicting ADHD (AUROC: 0.72). (25) Analyzing ∼6,000 candidate multimodal predictors, we found that the onset of ADHD in early adolescence was robustly (accuracy: ∼86%, AUROC: 0.919) predicted by a simple model comprising the presence of a disorder of excessive somnolence, two metrics of poor school performance and parent-child conflict.

Sleep disturbances are widely reported in ADHD including longer sleep latency, frequent awakenings, non-restorative sleep, decreased sleep and daytime somnolence (20, 26) Though many children with ADHD are treated with stimulants, the evidence that this disrupts sleep is inconclusive, though sleep disturbances are thought to worsen neurocognitive outcomes. (27) In the present study we included many types of sleep disorders and metrics as candidate predictors (**Supplementary Table 1**) and identified excessive somnolence as the most important predictor of the future onset of ADHD at 11-12 yrs when measured in children ages 9-10 yrs who have not been diagnosed with ADHD or taken stimulants. Thus, our findings extend prior work by suggesting that excessive daytime somnolence rather than other sleep metrics may be a helpful predictor of future ADHD case status. Excessive somnolence could be caused by a variety of developmental or environmental factors in school age children and future work may provide a mechanistic explanation of how it predicts ADHD onset. As noted above, poor school performance and family dysfunction have previously been identified as a predictor of ADHD and is well-associated with the disorder. Here, we add to this literature by identifying poor school performance and parent-child conflict as prospective predictors of ADHD onset in early adolescence. Of note, final predictors of new onset ADHD were essentially a subset of those that predicted all prevailing cases, where in the latter parent behavioral traits of total and externalizing problem behaviors were also present.

We found that prospective prediction of ADHD onset in early adolescence was not improved by neural features. However, our neural-only model of ADHD onset did achieve moderately strong performance (accuracy: ∼79%, AUROC: 0.841) and is of interest. The neural substrate of ADHD has been extensively studied in group-level associative work. More recently, the construction of ML classifiers with neural features was stimulated by the formation of the aggregated ADHD-200 dataset and associated Global Competition, though many resultant studies have been criticized for reporting “inflated” performance statistics based on cross-validated training rather than testing for generalization in held-out, unseen data. (28) Among the latter, performance has varied widely with accuracy rarely surpassing 80% and the majority of studies analyzing ADHD-200 cross-sectional, data with a wide age span. We are not aware of other studies using ML for prospective prediction of the onset of adolescent ADHD using a comparably large number of neural features across multiple MRI types in a standardized cohort. In new onset cases, we found the most prominent predictor was the correlation between the ventral attention network and right ventral diencephalon ROI, followed by SST contrast in the left pars opercularis (Brodmann Area 44) and cortical thickness in the right transverse temporal ROI (linked with the processing of incoming auditory information). The ventral attention network is one of the primary attention networks in the brain and directs attention to unexpected stimuli. It has been very well-associated with ADHD symptomatology in both children and adults, as have differences in subcortical structures. (29, 30, 31, 32, 33) Among subcortical structures, the diencephalon was historically less studied in ADHD. However, the thalamus, a primary component of the diencephalon which modulates and filters interfering stimuli has recently attracted much attention with structural thalamic differences identified in youth with ADHD. (34, 35, 36)

### Predicting the onset of Oppositional Defiant Disorder in adolescence

Oppositional defiant disorder is characterized by a pattern of uncooperative, defiant and angry behavior towards authority figures that causes significant problems at home or school. Like ADHD, a proportion of youth with ODD ‘grow out’ of the condition and ∼50% of youth with ODD have ADHD. Of those in whom ODD persists, CD may evolve and in adulthood ∼40% go on to develop antisocial personality disorder and/or other mental health or substance abuse problems. Since the prevalence of ODD climbs markedly in elementary school and empirically-based treatment is available, identifying specific prospective predictors of the disorder in late childhood and early adolescence – particularly those that differentiate it from ADHD - is of considerable import. ODD and CD are often grouped as the ‘disruptive disorders’ and unfortunately few large-scale ML studies have approached ODD in isolation. To our knowledge, this study represents the first to analyze a large number of multimodal predictors including multiple types of neuroimaging to prospectively predict ODD as distinct from CD in early adolescence. We found that deep learning optimized with IEL prospectively predicted the onset of ODD with strong performance using multimodal features (accuracy: ∼97%, AUROC: 0.996) in held-out, unseen data. While sleep disorders were final predictors that ODD shared with ADHD, ODD had a more complex predictive model that additionally included several measures of parental mental health problems (either parent has depression i.e. nerves or nervous breakdown problem; parent externalizing and aggressive problems) but did not include the metrics of school performance that predicted ADHD onset. Indeed, the most important predictor was whether the child had already come to clinical psychiatric attention prior to age 9-10 yrs. Here, our work is concordant with extant group-based studies in ODD, which associate case status with stress and conflict, parental depression and other parental factors such as hostility, support, scaffolding and further suggest that symptoms are present in preschool and ‘cascade’ toward eventual diagnosis with parental mental health problems significantly moderating treatment outcome. (37, 38, 39, 40, 41)

As with ADHD, we found that biological and physiologic metrics were not selected in multimodal prospective prediction of the onset of ODD, and that neural-only models sacrificed substantial performance. However, our neural-only model still obtained moderately strong performance (accuracy: 74%, AUROC: 0.792) and a striking result worthy of examination. While 5,777 neural features across multiple neuroimaging types were analyzed, ODD onset at 11-12 yrs was predicted by a markedly homogenous combination of features that were all rsfMRI metrics representing connectivity between cortical networks and subcortical ROIs, in particular limbic regions of the amygdala and caudate and putamen (dorsal striatum). Limbic regions in the left hemisphere predicted case status while those in the right hemisphere had an inverse relationship with ODD. Moreover, cortical networks selected as final predictors had intuitive relationships with ODD symptomatology, being associated with navigating and integrating learned social rules, hierarchies, and contingencies (salience); empathy and introspection (default); efficient task switching (cinguloopercular); and executive control (fronto-parietal). (42, 43, 44, 45, 46, 47, 48) Many have known limbic nodes where the latter structures are associated with fear and threat detection and the autonomic “fight or flight” response (amygdala) reinforcement learning and action selection (dorsal striatum). (49, 50, 51) As noted above, few large-scale ML predictive studies have focused exclusively on ODD or made head-to-head comparisons among the externalizing disorders, including neuroimaging studies. Menon & Krishnamurthy predicted disruptive behaviors (collapsing ODD and CD into one category) in children age 9-10 in the ABCD cohort using a convolutional neural network applied to three types of neural features (diffusion, structural and seed-based rsfMRI connectivity) obtained at 9-10 yrs to examine the relative predictive power of each type of imaging. They obtained moderate performance (accuracy: 0.72, AUROC: 0.74) without testing in held-out data and found that a combination of modalities performed better than any single imaging type. The right superior longitudinal fasciculus, middle frontal, postcentral, middle occipital and middle temporal gyri and inferior parietal lobule were class discriminative in disruptive behaviors. Thus, the current study suggests an intriguing jumping-off point for neural prediction of ODD development in suggesting a focus on cortical-subcortical relationships centered around connectivity between cortical control networks and limbic loci performing emotional response and action selection. In this, ODD contrasts with the attention and language processing networks and areas that were emphasized in ADHD onset.

### Predicting the onset of Conduct Disorder in early adolescence

While conduct disorder may be grouped with ODD as the ‘disruptive’ disorders, it is differentiated by the presence of aggression and destructive behaviors directed toward people, animals or property, serious violation of rules and lack of empathy. CD is often considered the most severe and disabling of the adolescent externalizing disorders. While ∼60-70% of youth will lose the diagnosis in adulthood, those that do not have a relatively poor prognosis associated with the development of other mental health and substance abuse disorders, antisocial personality disorder and life impairment including involvement in the justice system. In prior group-based longitudinal studies, the development of CD has been associated with impulsivity, parental behaviors such as poor supervision and punitive discipline and cold or antisocial parental traits and parental conflict, family risk factors like large size or low income and contextual factors such as antisocial peers, and adverse school or neighborhood environments. (52) In the only multimodal ML classification study previously performed in CD specifically, Chan et al also used data collected from children age 9-10 yrs in the ABCD cohort to predict all prevailing cases of CD at 11-12 yrs. (14) This study employed artificial neural networks and 52 candidate predictors comprising 20 graph metrics computed from rsfMRI, 16 psychosocial features selected empirically based on prior literature, 4 basic demographic descriptors and 9 cognitive metrics derived from psychometrics testing. In contrast to the present study, CD, ADHD and ODD symptomatology at 9-10 yrs were also allowed as candidate predictors. This design achieved 91% accuracy and 0.96 AUROC compared to our 96% accuracy and 0.99 AUROC in prospective prediction of all prevailing cases of CD at 11-12 yrs. They found that greater ADHD and ODD symptomatology, frontoparietal efficiency and reports of family members throwing objects predicted future CD while lower crystallized cognitive and card sorting ability, subcortical efficiency, frontoparietal degree, family income and parental monitoring were inversely related to case status. This study is comparable to our multimodal model predicting all prevailing cases at 11-12 yrs, though we analyzed a larger number of candidate predictors and more types of neural metrics. We similarly found parent-child conflict to be an important final predictor, but otherwise final predictors emphasized sleep disturbance (sleep-wake transition), poor school performance, parent somatization and aggressive traits and structural brain differences in the left transverse temporal and anterior cingulate cortex (ACC) ROIs.

When focusing on predicting new onset cases of CD with multimodal features we identified a more parsimonious model with strong predictive performance (accuracy: 90%, AUROC: 0.922) where family conflict, sleep disturbances (sleep-wake transition disorder and total disturbances) and whether the child had come to clinical attention prior to 9-10 yrs were important predictors. It is notable that among the three adolescent externalizing disorders, CD is the only condition in which neural predictors were selected as final predictors among ∼6,000 multimodal candidate predictors after extensive AI-guided feature selection. In prospectively predicting the onset of CD, structural differences in the left hippocampus ROI interacted with psychosocial factors to drive prediction of case status. Prior group-based studies (including in the ABCD cohort) have identified associations between CD symptomatology and structural and functional differences in the limbic system (which includes the hippocampus), ACC, orbito-frontal, prefrontal and temporal cortices, though not all studies segregate CD from ODD. (53, 54, 55, 56) Extant work has also specifically identified aberrant volumes in paralimbic structures including hippocampal ROIs in incarcerated adults and youth with psychopathic traits. (57, 58, 59) Though the hippocampus is known for its role in memory formation, it is deeply interconnected with other limbic structures and plays a prominent role in fear conditioning and affective processes. (60)

While neural features proved more important in multimodal prediction of CD vs ADHD and ODD, when we restricted candidate predictors to only neural features, performance dropped substantially. Similarly, Chan et al found that accuracy dropped to 77% and AUROC to 80% when only neural features were used to make prospective predictions of all prevailing CD cases at 11-12 yrs, a very comparable performance differential. However, moderately strong performance was still obtained (accuracy: 80%, AUROC: 0.808) giving credence to these findings. In the neural-only model, features in good concordance with prior literature were identified, with differences in frontal, temporal and limbic (caudate, amygdala, hippocampus) structures and connectivity between the cinguloopercular network and amygdala appearing as important predictors of CD onset in early adolescence. While some regional ROIs, particularly limbic structures, were common to ODD and CD, we found that neural predictors of CD onset emphasized structural over connectivity features and the hippocampus appeared as limbic structural predictor that was specific to CD.

## CONCLUSION

Taken together, our results suggest that highly accurate (>85%) prediction of the onset of each of the early adolescent externalizing disorders is possible using ML optimized with AI and that individual-level prospective prediction of ADHD, ODD and CD benefits from the inclusion of multimodal features drawn from multiple knowledge domains, particularly psychosocial predictors related to sleep disorders, parent mental health and behavioral traits and school performance. In CD specifically - but not ADHD or ODD - metrics derived from structural MRI interacted with psychosocial features in predicting later case onset and these neural features and may hold particular promise in identifying children at risk for this highly disabling disorder. Cognitive features derived from psychometric testing and other forms of physiologic data (e.g. hormonal levels, biometric testing) were deemphasized throughout our experiments. Among neural features, metrics related to subcortical ROIs and connectivity between cortical and subcortical ROIs were prominent, congruent with the existing literature in externalizing disorders. In terms of MRI modalities, structural gray and white matter features and rsfMRI-derived connectivity were valuable prospective predictors across all three disorders, with tfMRI only appearing in ADHD and no diffusion MRI metrics featured. We achieved high performance across all multimodal experiments and identified a strong correlation between accuracy and final predictor importance, suggesting that automated feature selection with AI techniques such as IEL can facilitate the discovery of impactful predictors among high dimension data in a principled manner and generate robust predictive models.

## LIMITATIONS

This study uses secondary data from the ABCD study. We were therefore unable to control for any bias during data collection and there is a mild bias toward higher-income participant families of white race in the early adolescent cohort, though the ABCD study strived for population representation. Similarly, the externalizing disorders have shown differences in population case ascertainment associated with characteristics such as sex/gender, race/ethnicity and sociodemographic factors which have varied over time and are still the subject of ongoing research. We do not take a position on these phenomena in this study and constructed balanced samples based on case ascertainment using the CBCL. Cases were matched with controls based on natal sex and age. However, sex, gender, race, ethnicity and many sociodemographic and cultural factors were included as candidate predictors as is standard practice in large scale ML studies and which does allow for the influence of such factors to be revealed. Further work using a similar design in participant samples stratified by sex/gender or race/ethnicity could also elucidate differential effects. Data is not available prior to baseline (age 9-10 years) assessment and we cannot therefore conclusively rule out that youth participants may have met criteria based on the CBCL for ADHD, ODD or CD prior to ≤8 yrs. Thus, it is possible that certain cases coded as ‘new onset’ at 11-12 years of age could have met clinical criteria at ≤8 yrs but not at 9-10 yrs. In the present study, we defined cases as an individual meeting ASEBA clinical thresholds in CBCL subscale scores pertinent to ADHD, ODD and CD and did not exclude participants who thereby met criteria for other conditions. Thus, co-morbidity may be present in the experimental samples as is common in clinical populations and occurs in most research studies in early adolescence. Our study is not exhaustive. It is possible that different results could have been obtained if more or different candidate predictors were included. We tested for generalization in a holdout, unseen test set obtained by partitioning the data, a gold standard method in ML. However, methods and results should also be tested for replication in an external dataset other than ABCD.

## Data Availability

All data produced in the present work are contained in the manuscript

https://nda.nih.gov/abcd/abcd-annual-releases.html

## ACKNOWLEDGEMENTS

Research reported in this publication was supported by the National Institute of Mental Health of the National Institutes of Health under award number R00MH118359 to NdL. The content is solely the responsibility of the authors and does not necessarily represent the official views of the National Institutes of Health. The support and resources from the Center for High Performance Computing at the University of Utah are also gratefully acknowledged.

Data used in the preparation of this article were obtained from the Adolescent Brain Cognitive Development^SM^ (ABCD) Study (https://abcdstudy.org), held in the NIMH Data Archive (NDA). This is a multisite, longitudinal study designed to recruit more than 10,000 children age 9-10 and follow them over 10 years into early adulthood. The ABCD Study® is supported by the National Institutes of Health and additional federal partners under award numbers U01DA041048, U01DA050989, U01DA051016, U01DA041022, U01DA051018, U01DA051037, U01DA050987, U01DA041174, U01DA041106, U01DA041117, U01DA041028, U01DA041134, U01DA050988, U01DA051039, U01DA041156, U01DA041025, U01DA041120, U01DA051038, U01DA041148, U01DA041093, U01DA041089, U24DA041123, U24DA041147. Additional support for this work was made possible from NIEHS R01-ES032295 and R01-ES031074. A full list of supporters is available at https://abcdstudy.org/federal-partners.html. A listing of participating sites and a complete listing of the study investigators can be found at https://abcdstudy.org/consortium_members/. ABCD consortium investigators designed and implemented the study and/or provided data but did not necessarily participate in the analysis or writing of this report. This manuscript reflects the views of the authors and may not reflect the opinions or views of the NIH or ABCD consortium investigators. The ABCD data repository grows and changes over time. The ABCD data used in this report came from 10.15154/1523041. DOIs can be found at https://nda.nih.gov/abcd/abcd-annual-releases.html.

