## Supplementary Figure 1 for "Identifying selective predictors of ADHD, Oppositional Defiant and Conduct Disorder onset in early adolescence with optimized deep learning"

Receiver Operating Characteristic (ROC) curves are shown for multimodal and neural-only models of new onset cases of ADHD, Oppositional Defiant Disorder (ODD) and Conduct Disorder (CD) at 11-12 yrs predicted with features measured at 9-10 yrs.

|  | **Multimodal** | **Neural-only** |
| --- | --- | --- |
| **ADHD** | 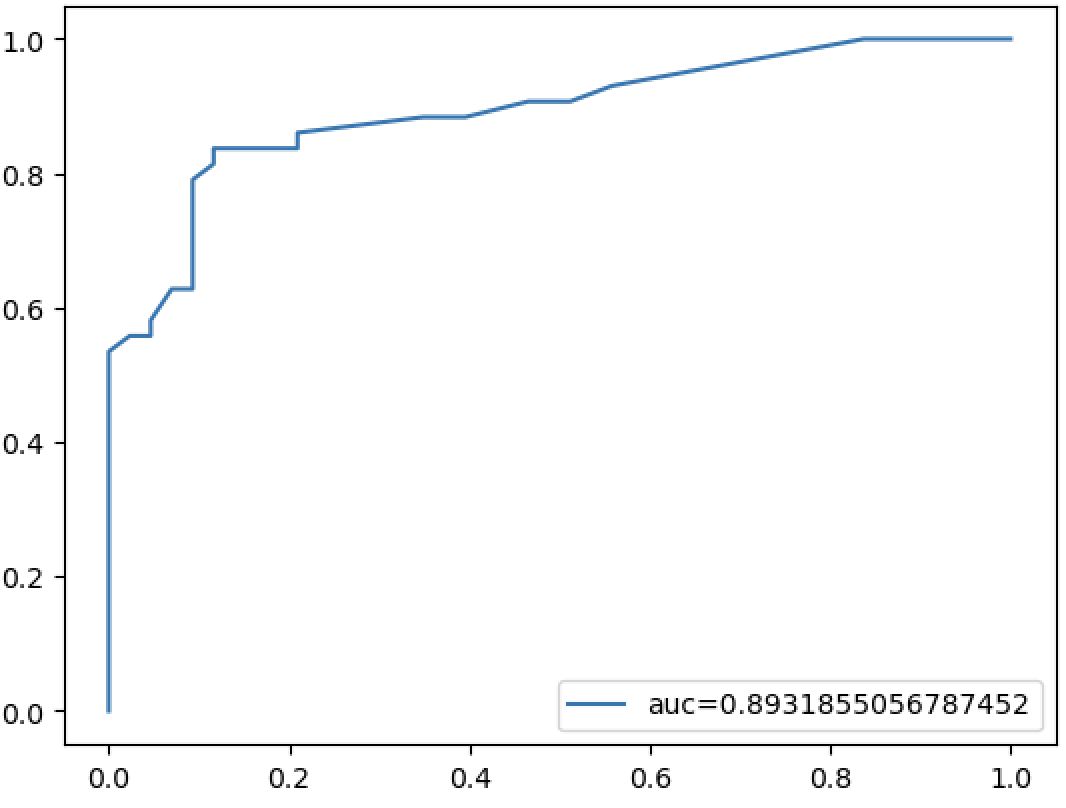 | 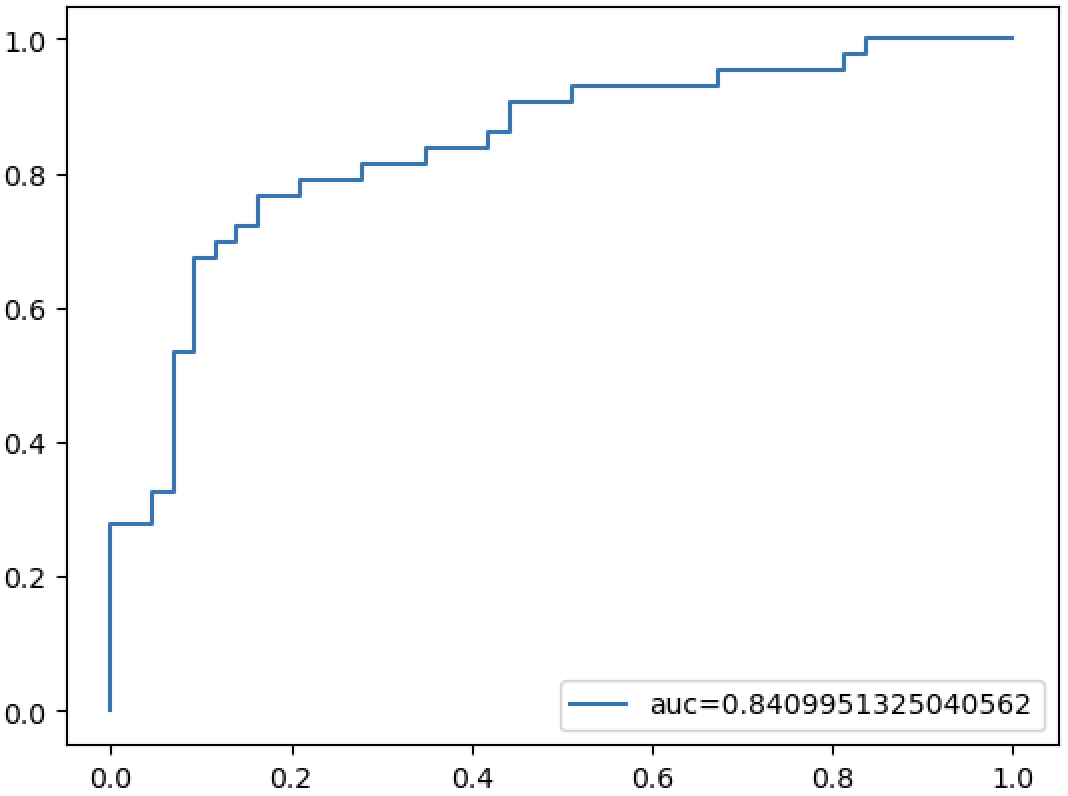 |
| **ODD** | 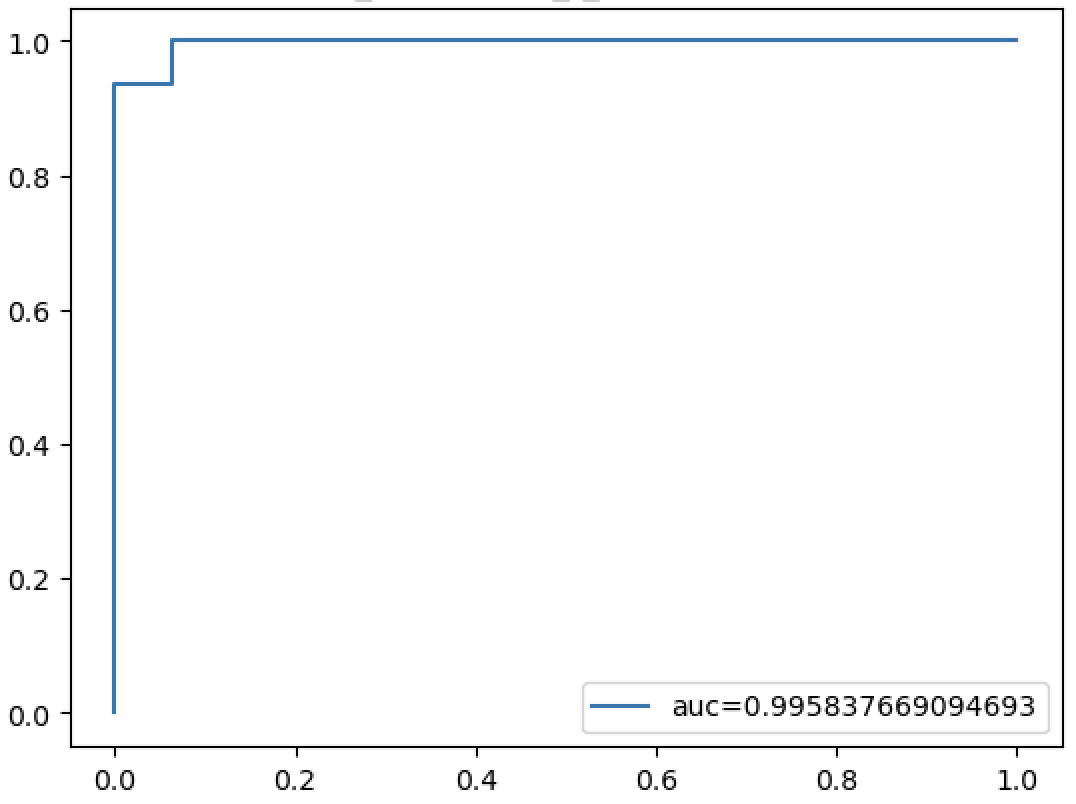 | 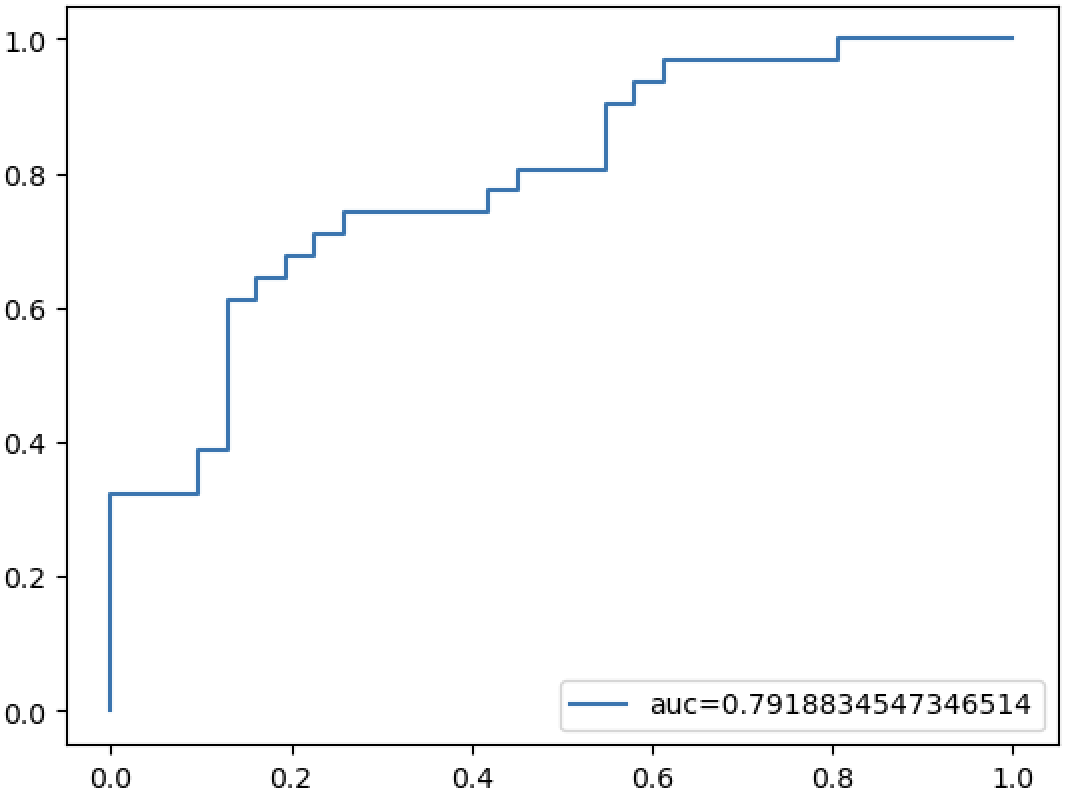 |
| **CD** | 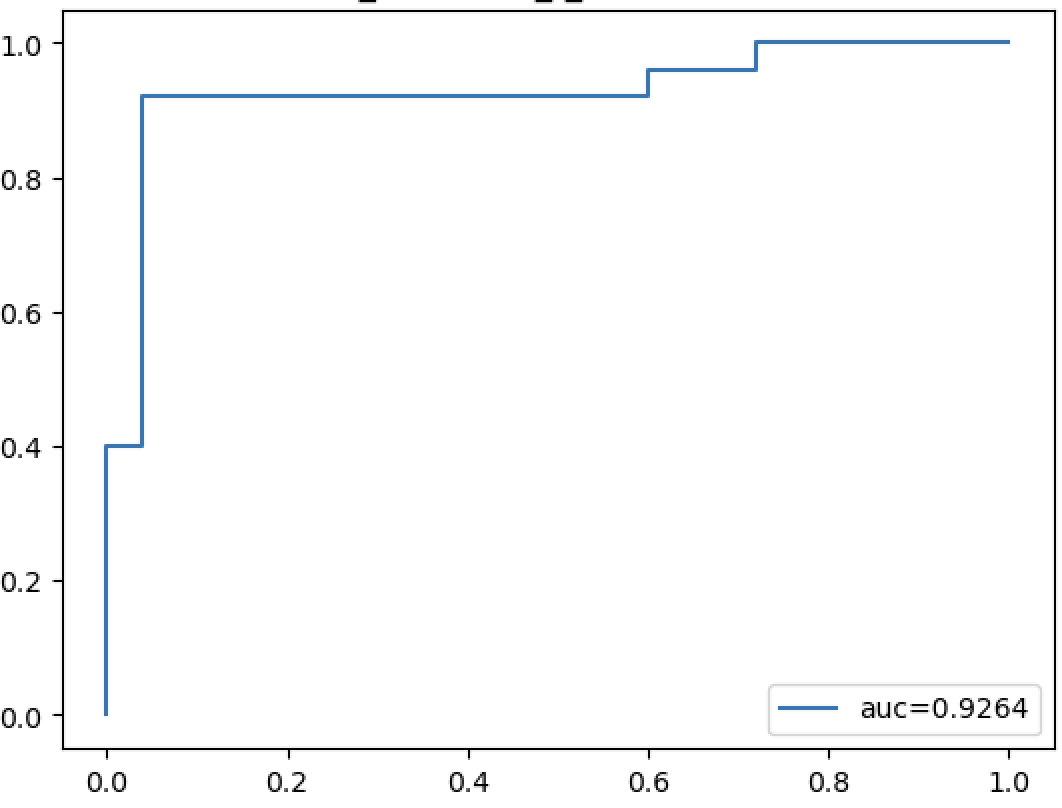 | 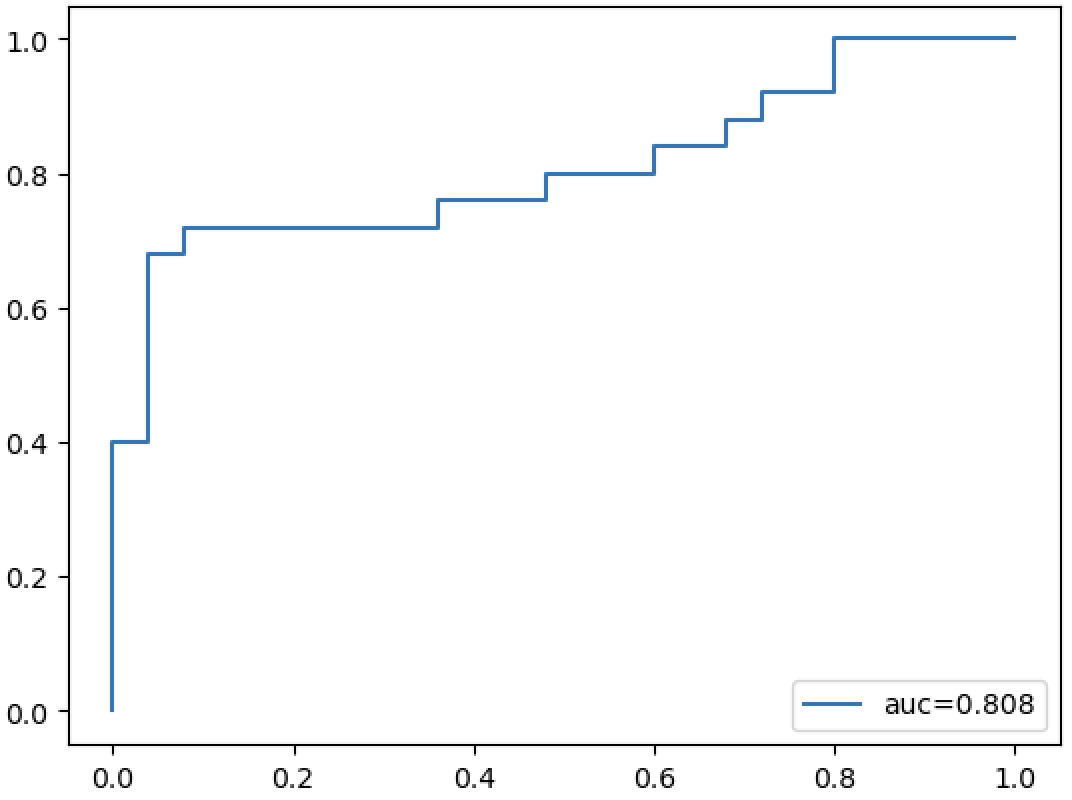 |
