## Supplementary Table 1 for "Identifying selective predictors of ADHD, Oppositional Defiant and Conduct Disorder onset in early adolescence with optimized deep learning"

**Supplementary Table X: Baseline ABCD Assessment Protocol**

We analyzed 5,777 input features from the ABCD assessment protocol collected from youth and their parents at baseline, when youth were 9-10 years of age. Instruments and imaging protocols from which features were extracted for inclusion in our selection pipeline are shown below. This list is adapted from the ABCD study protocol site which may be accessed at <https://abcdstudy.org/scientists/protocols/>.

**Youth Protocol**

**Physical Health**

PhenX Anthropometrics

Snellen Vision Screener

Edinburgh Handedness Inventory

Youth Risk Behavior Survey – Exercise

Pubertal Development Scale

Menstrual Cycle Survey

Screen Time Survey

**Brain Imaging**

Structural MRI

- 3D T1 – Weighted
- 3D T2 – Weighted
- Diffusion Tensor Imaging

Functional MRI

- Resting State
- Monetary Incentive Delay Task
- Stop Signal Task
- Emotional N-Back Task

Site

Motion statistics for each MRI modality

**Biospecimens**

Oral fluids: pubertal hormones

**Mental Health**

UPPS-P for Children – Short Form (ABCD version)

PhenX Behavioral Inhibition/Behavioral Approach System (BIS/BAS) Scales

Youth Resilience Scale

**Neurocognition**

NIH Toolbox Tasks:

- Picture Vocabulary
- Flanker Inhibitory Control & Attention
- List Sorting Working Memory
- Dimensional Change Card Sort
- Pattern Comparison Processing Speed
- Picture Sequence Memory
- Oral Reading Recognition

Rey Auditory Verbal Learning Task

Cash Choice Task

Little Man Task

Matrix Reasoning Task

RAVLT Delayed Recall

**Substance Use**

Participant Last Use Survey (PLUS) for substance use within the last 24 hours

PhenX Peer Group Deviance Survey

PATH Intention to Use Tobacco Survey

Timeline Follow-Back Survey

Caffeine Intake Survey

**Culture and Environment**

Prosocial Behavior Survey

PhenX Acculturation Survey

Parental Monitoring Survey

Acceptance Subscale from Children’s Report of Parental Behavior Inventory – Short

PhenX Family Environment Scale – Family Conflict

PhenX Neighborhood Safety/Crime Survey

PhenX School Risk & Protective Factors Survey

**Parent Protocol**

**Physical Health**

PhenX Demographics Survey

Medical History Questionnaire

Developmental History Questionnaire

PhenX Medications Survey

Pubertal Development Scale

Menstrual Cycle Survey

Sleep Disturbances Scale for Children

Sports and Activities Involvement Questionnaire

Screen Time Survey

Ohio State TBI Screen – Short

**Mental Health**

Adult Self Report Survey

Family History Assessment Survey

**Substance Use**

Participant Last Use Survey (PLUS) for substance use within the last 24 hours

Parent Rules Survey

PhenX Community Risk and Protective Factors

**Culture and Environment**

Vancouver Index of Acculturation – Short Survey

Multi-Group Ethnic Identity Measure – R Survey

Prosocial Behavior Survey

Mexican American Cultural Values Scale

PhenX Acculturation Survey

PhenX Family Environment Scale – Family Conflict

PhenX Neighborhood Safety/Crime Survey

Native American Acculturation Scale

**Other Data Sources**

Geocoding from Residential History

School Records

Brief Problem Monitor – Teacher Form
